# Development and internal validation of early- and mid-pregnancy prediction models for macrosomia in nulliparous singleton pregnancies

**DOI:** 10.64898/2026.10.29.26366704

**Authors:** Yanqi Wu, Myrthe van der Ven, Elisabetta Peri, Sima Asvadi, M. Beatrijs van der Hout-van der Jagt, S. Guid Oei, Massimo Mischi, Xi Long

## Abstract

**Purpose:** Macrosomia is associated with adverse maternal and neonatal outcomes, but early risk stratification remains challenging. Most existing prediction approaches rely on later-pregnancy ultrasound measurements, limiting opportunities for earlier preventive counseling and monitoring.

**Methods:** In this secondary analysis of the prospective nuMoM2b cohort in the United States, we included 6,371 nulliparous singleton pregnancies selected for available periconceptional diet and ultrasound data at Visits 2 and 3. We developed visit-updated multivariable logistic regression models for birthweight >4000 g and >4500 g using routinely available maternal characteristics, anthropometry, periconceptional diet, and serial ultrasound measurements, with sensitivity analyses using large-for-gestational-age birthweight. Model performance was assessed by repeated stratified cross-validation and compared with World Health Organization fetal growth chart-based ultrasound percentile benchmarks, emphasizing the estimated fetal weight percentile as the most clinically familiar proxy.

**Results:** Birthweight >4000 g and >4500 g occurred in 7.9% and 1.2% of pregnancies, respectively. Models updated with later pregnancy information achieved the best discrimination, with an area under the receiver operating characteristic curve of 0.73 for birthweight >4000 g and 0.82 for birthweight >4500 g. In this selected analytic cohort, the early-pregnancy model based on maternal data alone provided moderate discrimination (0.63 and 0.72, respectively), although lower than the strongest third-trimester WHO fetal growth chart-based ultrasound percentile benchmarks.

**Conclusion:** Within this selected cohort, routinely available early-pregnancy maternal characteristics and anthropometry provided moderate risk stratification for macrosomia, and later ultrasound measurements improved performance further. Estimates for birthweight >4500 g require caution given the 77 events. External validation is required before clinical implementation.

**What this study adds to the clinical work:** Routinely available early-pregnancy maternal characteristics and anthropometry provided moderate risk stratification for macrosomia in nulliparous singleton pregnancies. Visit-updated models incorporating later ultrasound measurements improve discrimination further and may help refine surveillance during pregnancy.

## 1. Introduction

Fetal macrosomia is associated with increased maternal and neonatal morbidity, including birth trauma and postpartum hemorrhage [1,2]. Its definition is not uniform across studies, with birthweight thresholds of >4000 g and >4500 g both commonly used to capture varying degrees of risk [3,4]. The condition remains a pressing clinical concern, with approximately 8% of neonates in the United States weighing over 4000 g at birth [5,6].

Early identification of pregnancies at high risk for macrosomia enables timely lifestyle interventions— such as dietary modification and gestational weight gain management—which can substantially reduce its incidence [7–9]. However, predictions relying mainly on later-pregnancy ultrasound provide less opportunity for early preventive action. Furthermore, clinical assessments often depend on sonographic estimated fetal weight, which becomes less reliable at the extremes of birthweight [10]. While early prediction approaches exist [11,12], a recent systematic review of 58 multivariable models revealed that most still depend predominantly on near-term ultrasound [13]. Only five models incorporated second-trimester or earlier ultrasound data together with routinely available maternal characteristics [14–18]. This limitation is particularly critical for nulliparous pregnancies, in which previous obstetric history is unavailable for risk stratification.

Therefore, in this secondary analysis of the prospective nuMoM2b cohort [19], we aimed to develop and internally validate visit-updated multivariable prediction models for macrosomia (>4000 g and >4500 g) in nulliparous singleton pregnancies. Specifically, we evaluated the baseline predictive value of early-pregnancy maternal characteristics and periconceptional diet, quantified the incremental benefit of second-and third-trimester ultrasound, and benchmarked model performance against World Health Organization (WHO) fetal growth chart percentiles [10,14–18].

## 2. Materials and methods

### 2.1 Study design and participants

This was a secondary analysis of the prospective Nulliparous Pregnancy Outcomes Study: Monitoring Mothers-to-Be (nuMoM2b) cohort [19]. Nulliparous women with singleton pregnancies were recruited between 2010 and 2013 from hospitals affiliated with eight clinical centers in the United States [19]. Three standardized prenatal visits were scheduled at 6–13 weeks (Visit 1), 16–21 weeks (Visit 2), and 22–29 weeks (Visit 3) of gestation [19]. The original study was approved by the institutional review board at each participating site, and all participants provided written informed consent before enrollment [19]. For the present analysis, pregnancies were excluded if periconceptional dietary data, ultrasound fetal biometry measurements from Visit 2 or Visit 3, birth outcome data, or live-birth status were unavailable. Candidate predictors are summarized in Table 1 and Table 2.

**Table 1.**
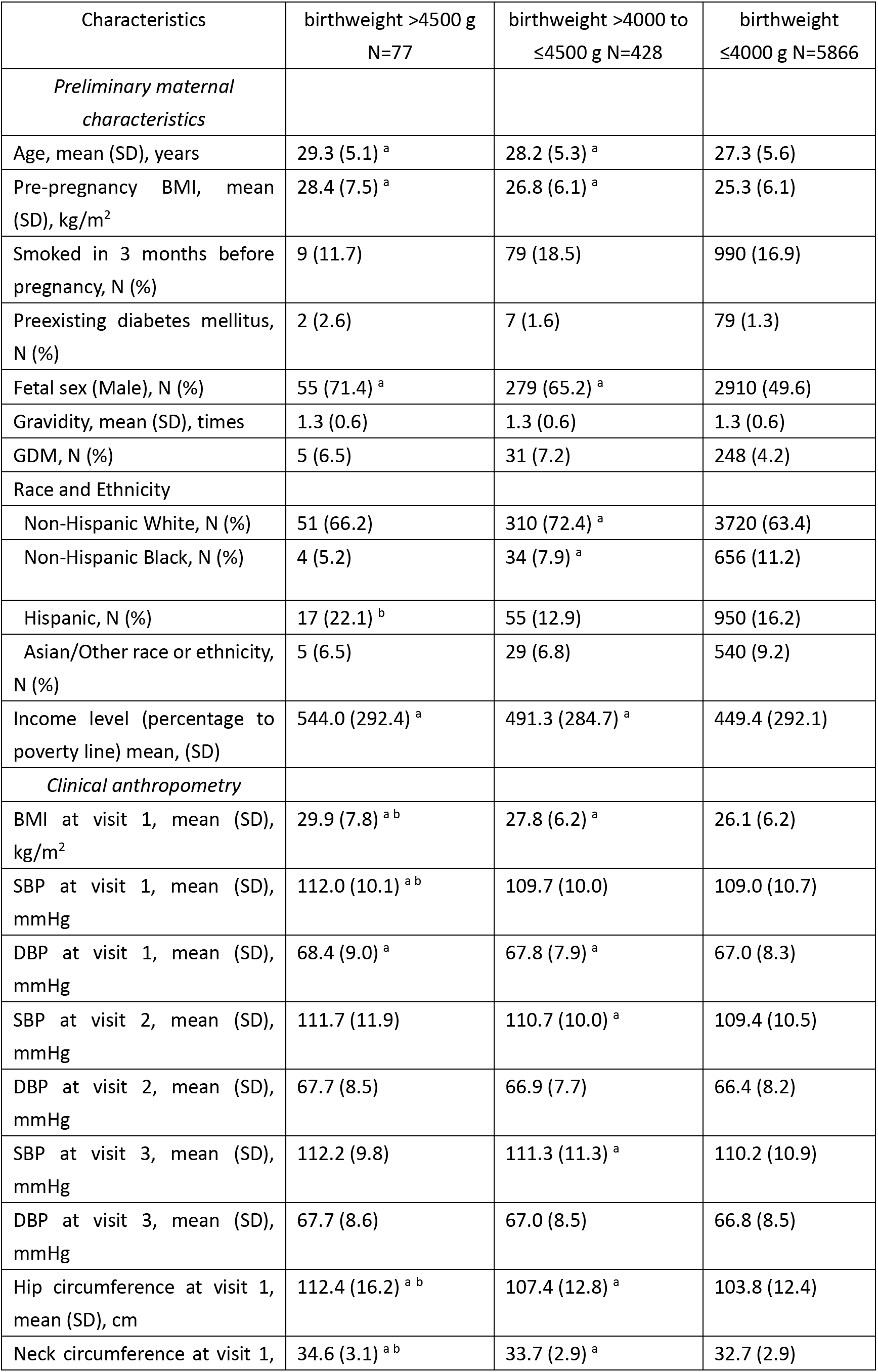
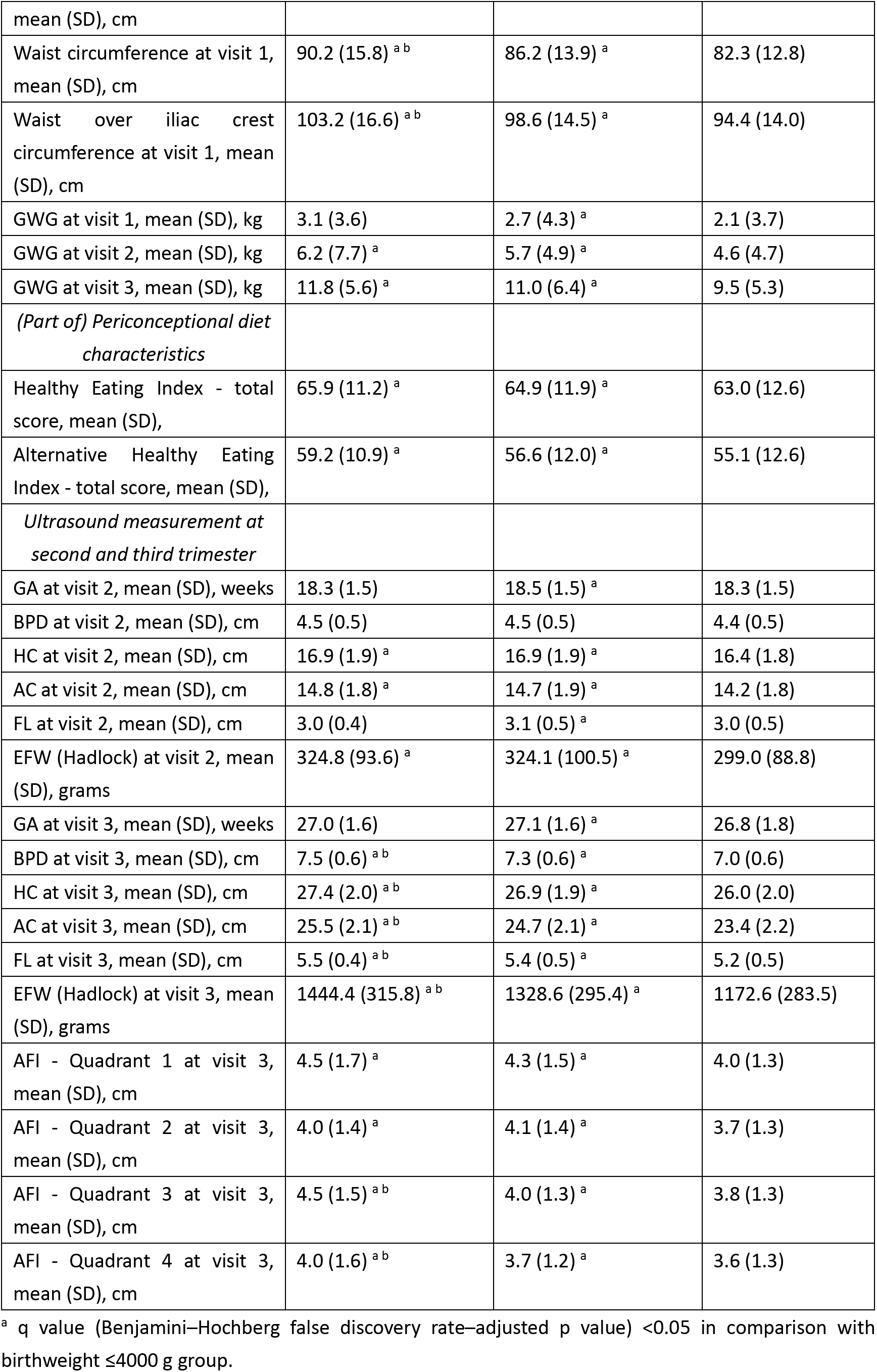
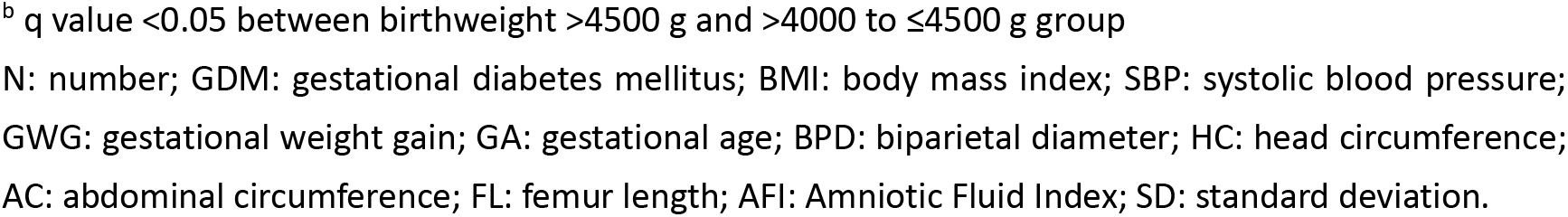
Pregnancy characteristics by birthweight category (>4500 g, >4000 to ≤4500 g, and ≤4000 g).

| Characteristics | birthweight >4500 g<br>N=77 | birthweight >4000 to<br>≤4500 g N=428 | birthweight<br>≤4000 g N=5866 |
| --- | --- | --- | --- |
| *Preliminary maternal characteristics* |  |  |  |
| Age, mean (SD), years | 29.3 (5.1) <sup>a</sup> | 28.2 (5.3) <sup>a</sup> | 27.3 (5.6) |
| Pre-pregnancy BMI, mean (SD), kg/m<sup>2</sup> | 28.4 (7.5) <sup>a</sup> | 26.8 (6.1) <sup>a</sup> | 25.3 (6.1) |
| Smoked in 3 months before pregnancy, N (%) | 9 (11.7) | 79 (18.5) | 990 (16.9) |
| Preexisting diabetes mellitus, N (%) | 2 (2.6) | 7 (1.6) | 79 (1.3) |
| Fetal sex (Male), N (%) | 55 (71.4) <sup>a</sup> | 279 (65.2) <sup>a</sup> | 2910 (49.6) |
| Gravidity, mean (SD), times | 1.3 (0.6) | 1.3 (0.6) | 1.3 (0.6) |
| GDM, N (%) | 5 (6.5) | 31 (7.2) | 248 (4.2) |
| Race and Ethnicity |  |  |  |
| Non-Hispanic White, N (%) | 51 (66.2) | 310 (72.4) <sup>a</sup> | 3720 (63.4) |
| Non-Hispanic Black, N (%) | 4 (5.2) | 34 (7.9) <sup>a</sup> | 656 (11.2) |
| Hispanic, N (%) | 17 (22.1) <sup>b</sup> | 55 (12.9) | 950 (16.2) |
| Asian/Other race or ethnicity, N (%) | 5 (6.5) | 29 (6.8) | 540 (9.2) |
| Income level (percentage to poverty line) mean, (SD) | 544.0 (292.4) <sup>a</sup> | 491.3 (284.7) <sup>a</sup> | 449.4 (292.1) |
| *Clinical anthropometry* |  |  |  |
| BMI at visit 1, mean (SD), kg/m<sup>2</sup> | 29.9 (7.8) <sup>a b</sup> | 27.8 (6.2) <sup>a</sup> | 26.1 (6.2) |
| SBP at visit 1, mean (SD), mmHg | 112.0 (10.1) <sup>a b</sup> | 109.7 (10.0) | 109.0 (10.7) |
| DBP at visit 1, mean (SD), mmHg | 68.4 (9.0) <sup>a</sup> | 67.8 (7.9) <sup>a</sup> | 67.0 (8.3) |
| SBP at visit 2, mean (SD), mmHg | 111.7 (11.9) | 110.7 (10.0) <sup>a</sup> | 109.4 (10.5) |
| DBP at visit 2, mean (SD), mmHg | 67.7 (8.5) | 66.9 (7.7) | 66.4 (8.2) |
| SBP at visit 3, mean (SD), mmHg | 112.2 (9.8) | 111.3 (11.3) <sup>a</sup> | 110.2 (10.9) |
| DBP at visit 3, mean (SD), mmHg | 67.7 (8.6) | 67.0 (8.5) | 66.8 (8.5) |
| Hip circumference at visit 1, mean (SD), cm | 112.4 (16.2) <sup>a b</sup> | 107.4 (12.8) <sup>a</sup> | 103.8 (12.4) |
| Neck circumference at visit 1, | 34.6 (3.1) <sup>a b</sup> | 33.7 (2.9) <sup>a</sup> | 32.7 (2.9) |
| mean (SD), cm |  |  |  |
| Waist circumference at visit 1,<br>mean (SD), cm | 90.2 (15.8)<sup>a b</sup> | 86.2 (13.9)<sup>a</sup> | 82.3 (12.8) |
| Waist over iliac crest<br>circumference at visit 1, mean<br>(SD), cm | 103.2 (16.6)<sup>a b</sup> | 98.6 (14.5)<sup>a</sup> | 94.4 (14.0) |
| GWG at visit 1, mean (SD), kg | 3.1 (3.6) | 2.7 (4.3)<sup>a</sup> | 2.1 (3.7) |
| GWG at visit 2, mean (SD), kg | 6.2 (7.7)<sup>a</sup> | 5.7 (4.9)<sup>a</sup> | 4.6 (4.7) |
| GWG at visit 3, mean (SD), kg | 11.8 (5.6)<sup>a</sup> | 11.0 (6.4)<sup>a</sup> | 9.5 (5.3) |
| *(Part of) Periconceptional diet*<br>*characteristics* |  |  |  |
| Healthy Eating Index - total<br>score, mean (SD), | 65.9 (11.2)<sup>a</sup> | 64.9 (11.9)<sup>a</sup> | 63.0 (12.6) |
| Alternative Healthy Eating<br>Index - total score, mean (SD), | 59.2 (10.9)<sup>a</sup> | 56.6 (12.0)<sup>a</sup> | 55.1 (12.6) |
| *Ultrasound measurement at*<br>*second and third trimester* |  |  |  |
| GA at visit 2, mean (SD), weeks | 18.3 (1.5) | 18.5 (1.5)<sup>a</sup> | 18.3 (1.5) |
| BPD at visit 2, mean (SD), cm | 4.5 (0.5) | 4.5 (0.5) | 4.4 (0.5) |
| HC at visit 2, mean (SD), cm | 16.9 (1.9)<sup>a</sup> | 16.9 (1.9)<sup>a</sup> | 16.4 (1.8) |
| AC at visit 2, mean (SD), cm | 14.8 (1.8)<sup>a</sup> | 14.7 (1.9)<sup>a</sup> | 14.2 (1.8) |
| FL at visit 2, mean (SD), cm | 3.0 (0.4) | 3.1 (0.5)<sup>a</sup> | 3.0 (0.5) |
| EFW (Hadlock) at visit 2, mean<br>(SD), grams | 324.8 (93.6)<sup>a</sup> | 324.1 (100.5)<sup>a</sup> | 299.0 (88.8) |
| GA at visit 3, mean (SD), weeks | 27.0 (1.6) | 27.1 (1.6)<sup>a</sup> | 26.8 (1.8) |
| BPD at visit 3, mean (SD), cm | 7.5 (0.6)<sup>a b</sup> | 7.3 (0.6)<sup>a</sup> | 7.0 (0.6) |
| HC at visit 3, mean (SD), cm | 27.4 (2.0)<sup>a b</sup> | 26.9 (1.9)<sup>a</sup> | 26.0 (2.0) |
| AC at visit 3, mean (SD), cm | 25.5 (2.1)<sup>a b</sup> | 24.7 (2.1)<sup>a</sup> | 23.4 (2.2) |
| FL at visit 3, mean (SD), cm | 5.5 (0.4)<sup>a b</sup> | 5.4 (0.5)<sup>a</sup> | 5.2 (0.5) |
| EFW (Hadlock) at visit 3, mean<br>(SD), grams | 1444.4 (315.8)<sup>a b</sup> | 1328.6 (295.4)<sup>a</sup> | 1172.6 (283.5) |
| AFI - Quadrant 1 at visit 3,<br>mean (SD), cm | 4.5 (1.7)<sup>a</sup> | 4.3 (1.5)<sup>a</sup> | 4.0 (1.3) |
| AFI - Quadrant 2 at visit 3,<br>mean (SD), cm | 4.0 (1.4)<sup>a</sup> | 4.1 (1.4)<sup>a</sup> | 3.7 (1.3) |
| AFI - Quadrant 3 at visit 3,<br>mean (SD), cm | 4.5 (1.5)<sup>a b</sup> | 4.0 (1.3)<sup>a</sup> | 3.8 (1.3) |
| AFI - Quadrant 4 at visit 3,<br>mean (SD), cm | 4.0 (1.6)<sup>a b</sup> | 3.7 (1.2)<sup>a</sup> | 3.6 (1.3) |

<sup>a</sup> q value (Benjamini–Hochberg false discovery rate–adjusted p value) <0.05 in comparison with birthweight ≤4000 g group.
<sup>b</sup> q value <0.05 between birthweight >4500 g and >4000 to ≤4500 g group N: number; GDM: gestational diabetes mellitus; BMI: body mass index; SBP: systolic blood pressure; GWG: gestational weight gain; GA: gestational age; BPD: biparietal diameter; HC: head circumference; AC: abdominal circumference; FL: femur length; AFI: Amniotic Fluid Index; SD: standard deviation.

**Table 2.**
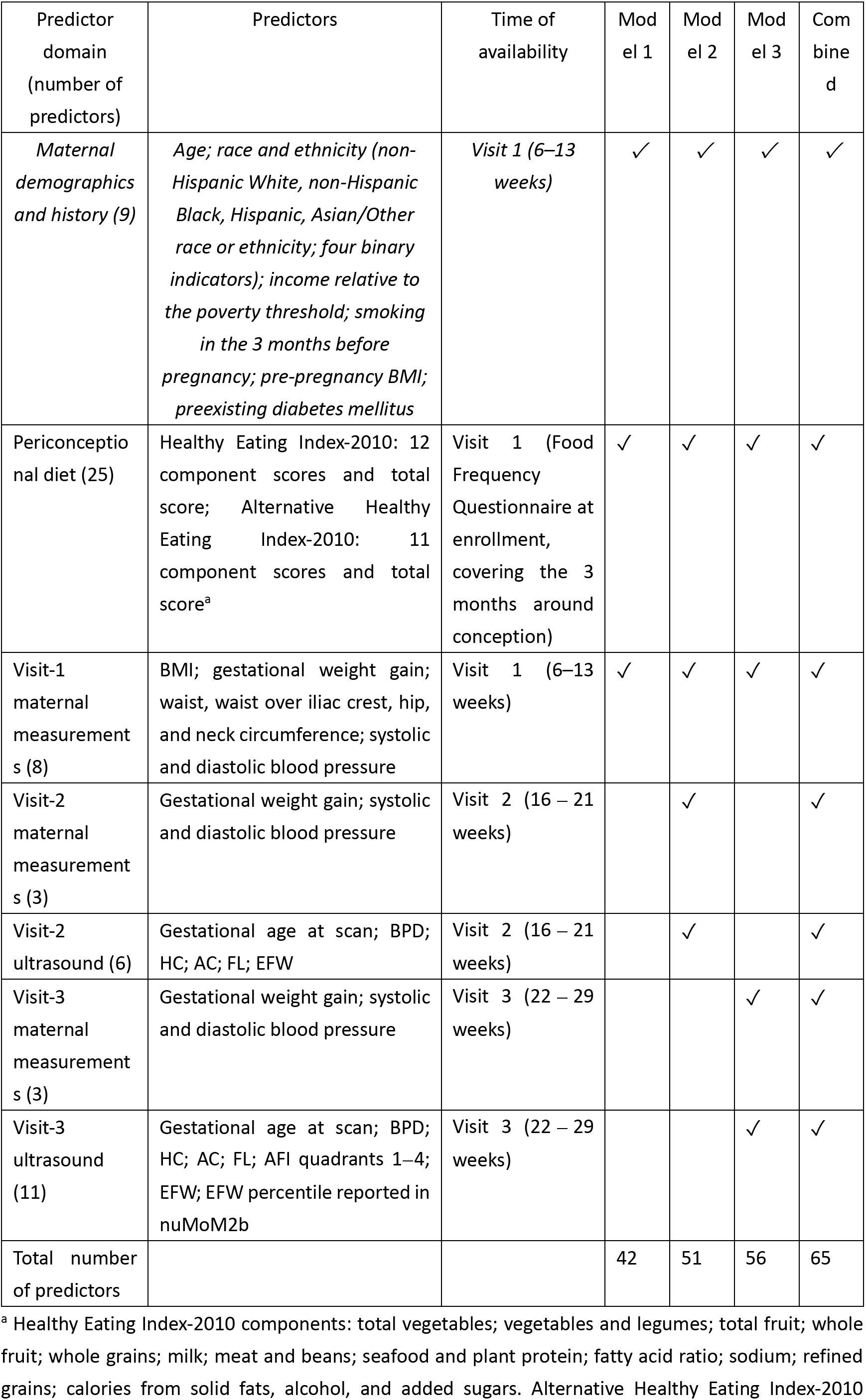
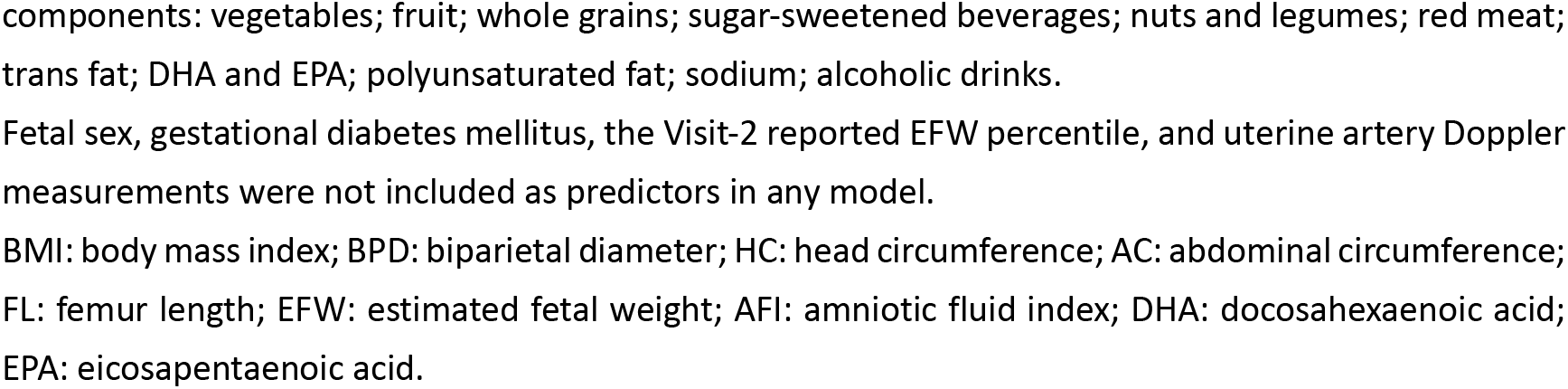
Predictors included in each prediction model and their time of availability.

| Predictor domain (number of predictors) | Predictors | Time of availability | Model 1 | Model 2 | Model 3 | Combined |
| --- | --- | --- | --- | --- | --- | --- |
| *Maternal demographics and history (9)* | *Age; race and ethnicity (non-Hispanic White, non-Hispanic Black, Hispanic, Asian/Other race or ethnicity; four binary indicators); income relative to the poverty threshold; smoking in the 3 months before pregnancy; pre-pregnancy BMI; preexisting diabetes mellitus* | *Visit 1 (6–13 weeks)* | ✓ | ✓ | ✓ | ✓ |
| Periconceptual diet (25) | Healthy Eating Index-2010: 12 component scores and total score; Alternative Healthy Eating Index-2010: 11 component scores and total score<sup>a</sup> | Visit 1 (Food Frequency Questionnaire at enrollment, covering the 3 months around conception) | ✓ | ✓ | ✓ | ✓ |
| Visit-1 maternal measurements (8) | BMI; gestational weight gain; waist, waist over iliac crest, hip, and neck circumference; systolic and diastolic blood pressure | Visit 1 (6–13 weeks) | ✓ | ✓ | ✓ | ✓ |
| Visit-2 maternal measurements (3) | Gestational weight gain; systolic and diastolic blood pressure | Visit 2 (16 – 21 weeks) |  | ✓ |  | ✓ |
| Visit-2 ultrasound (6) | Gestational age at scan; BPD; HC; AC; FL; EFW | Visit 2 (16 – 21 weeks) |  | ✓ |  | ✓ |
| Visit-3 maternal measurements (3) | Gestational weight gain; systolic and diastolic blood pressure | Visit 3 (22 – 29 weeks) |  |  | ✓ | ✓ |
| Visit-3 ultrasound (11) | Gestational age at scan; BPD; HC; AC; FL; AFI quadrants 1–4; EFW; EFW percentile reported in nuMoM2b | Visit 3 (22 – 29 weeks) |  |  | ✓ | ✓ |
| Total number of predictors |  |  | 42 | 51 | 56 | 65 |

<sup>a</sup> Healthy Eating Index-2010 components: total vegetables; vegetables and legumes; total fruit; whole fruit; whole grains; milk; meat and beans; seafood and plant protein; fatty acid ratio; sodium; refined grains; calories from solid fats, alcohol, and added sugars. Alternative Healthy Eating Index-2010
components: vegetables; fruit; whole grains; sugar-sweetened beverages; nuts and legumes; red meat; trans fat; DHA and EPA; polyunsaturated fat; sodium; alcoholic drinks. Fetal sex, gestational diabetes mellitus, the Visit-2 reported EFW percentile, and uterine artery Doppler measurements were not included as predictors in any model. BMI: body mass index; BPD: biparietal diameter; HC: head circumference; AC: abdominal circumference; FL: femur length; EFW: estimated fetal weight; AFI: amniotic fluid index; DHA: docosahexaenoic acid; EPA: eicosapentaenoic acid.

### 2.2 Outcomes

The primary outcomes were birthweight >4000 g and >4500 g. These thresholds were selected because they are widely used absolute definitions of macrosomia and represent clinically relevant degrees of fetal overgrowth. Separate prediction models were developed for each threshold.

Because absolute birthweight thresholds do not account for gestational age at delivery, we additionally performed sensitivity analyses using large-for-gestational-age birthweight. LGA90 and LGA97 were defined as birthweight above the 90th and 97th percentiles, respectively, for gestational age at delivery and neonatal sex according to the INTERGROWTH-21st Newborn Size Standards [20]. These analyses assessed whether the visit-updated prediction pattern was consistent when fetal overgrowth was defined relative to gestational age rather than by fixed birthweight thresholds.

### 2.3 Candidate predictors

Candidate predictors were prespecified according to clinical availability during antenatal care and prior evidence linking these factors to fetal overgrowth. Early-pregnancy maternal characteristics included age, pre-pregnancy body mass index, income level relative to the state-specific poverty threshold, race and ethnicity, smoking during the 3 months before pregnancy, and preexisting diabetes mellitus. Visit-specific maternal anthropometric measurements included body mass index, gestational weight gain, blood pressure, circumference measurements at Visit 1, and gestational weight gain and blood pressure at Visits 2 and 3. Fetal sex was used to define the LGA outcomes and in descriptive and univariate analyses; it was excluded from prediction models because its time of ascertainment was not documented.

Periconceptional diet was assessed at enrollment using the modified Block 2005 Food Frequency Questionnaire, which captured usual dietary intake during the 3 months around conception [21]. Overall diet quality was summarized using the Healthy Eating Index-2010 and the Alternative Healthy Eating Index-2010 [22–24]. The component and total scores of both indices were used as dietary predictors. Ultrasound predictors were obtained at Visits 2 and 3. Visit 2 included gestational age at scan, biparietal diameter, head circumference, abdominal circumference, femur length, and estimated fetal weight. Visit 3 included the same biometry variables together with the four amniotic fluid index quadrants and the estimated fetal weight percentile reported in nuMoM2b. Estimated fetal weight was calculated using the Hadlock formula [25]. Optional uterine artery Doppler measurements were not included because of the high proportion of missing data. Ultrasound availability for cohort derivation was defined primarily using Visit 2 and Visit 3 fetal biometry, because these measurements directly reflect fetal growth. Residual missing values in retained predictors were handled by imputation as described in Section 2.5. Predictors were entered as continuous or binary variables, with race and ethnicity coded as four binary indicators; the predictors included in each model and their time of availability are listed in Table 2.

### 2.4 Fetal growth chart comparator

As a clinical benchmark, we implemented the trimester-specific World Health Organization fetal growth chart [26,27]. Percentiles were computed from nuMoM2b fetal biometry or estimated fetal weight together with gestational age at ultrasound. For pregnancies with ultrasound examinations at Visit 2 and Visit 3, the WHO chart provided five percentile estimates at Visit 2 and five additional percentile estimates at Visit 3, based on head circumference, biparietal diameter, abdominal circumference, femur length, and estimated fetal weight. All WHO-derived percentile variables were evaluated as ultrasound-based benchmark comparators in the discrimination analyses; these comparisons with the prediction models were descriptive. Because estimated fetal weight is the most clinically familiar summary measure of fetal size, the estimated fetal weight percentile was additionally highlighted as the most practice-oriented WHO chart-based indicator.

As a supplementary comparison, we also evaluated INTERGROWTH-21st fetal ultrasound percentiles within the same chart-based comparator framework [28,29]. These percentiles were assessed analogously to the WHO-derived percentiles as single-predictor ultrasound comparators for subsequent birthweight >4000 g and >4500 g. These chart-based percentiles describe fetal size at the time of ultrasound examination and do not directly project expected birthweight at delivery. Because the INTERGROWTH-21st standard for Hadlock-based estimated fetal weight applies from 18 weeks of gestation, the Visit-2 estimated fetal weight percentile could not be derived for scans performed before 18 weeks. Details of percentile computation and variable coding are provided in the Supplementary Methods and Supplementary Table S3 and Supplementary Table S10.

### 2.5 Model development and internal validation

We developed four visit-updated multivariable logistic regression models to reflect information availability across routine antenatal care (Table 2). Model 1 used maternal demographics and history, Visit-1 anthropometry, early gestational weight gain, and periconceptional diet indices (42 predictors). Model 2 extended Model 1 by adding Visit-2 anthropometry and second-trimester ultrasound biometry with derived estimated fetal weight (51 predictors). Model 3 extended Model 1 by adding Visit-3 anthropometry and third-trimester ultrasound measurements (56 predictors). The Combined model included all information from Visit 1, Visit 2, and Visit 3 (65 predictors). All four models were developed and evaluated in the same analytic cohort to allow direct comparison.

Logistic regression was selected as the primary modeling approach because of its interpretability, reproducibility, and suitability for clinically deployable risk prediction [30]. Predictor-level missingness before imputation was summarized in Supplementary Table S9. Missing data among retained candidate predictors were imputed using a k-nearest-neighbor (k=5) approach. To prevent information leakage, all preprocessing steps, including imputation and scaling, were performed after data splitting and within the training data only. Models were developed and internally validated using repeated stratified 10-fold cross-validation with 1000 repetitions [31]. The models used an elastic-net penalty, with regularization strength (C) and L1 ratio tuned within the training data only, using inner 3-fold cross-validation optimizing average precision; the search spaces are provided in the note to Supplementary Table S8. In each repetition, every participant received one out-of-fold predicted probability; these predictions were averaged across repetitions at the participant level, so that each participant contributed a single prediction to the evaluation. AUROC and AUPRC were calculated from these participant-level predictions, and 95% confidence intervals were obtained from 1,000 bootstrap resamples of participants. Differences in AUROC between models were estimated using paired bootstrap resampling of the same participants.

Class imbalance was handled using weighted logistic regression, with class weights computed within each cross-validation training fold. The negative class was assigned a weight of 1, and the positive class was weighted by the square root of the negative-to-positive sample ratio to avoid excessive up-weighting of rare positive cases. Because class weighting changes the relationship between model scores and absolute outcome probabilities, predicted probabilities were then recalibrated using Platt scaling, with the Platt calibrator fitted within the training folds only and applied to the held-out fold. For the main model evaluations, calibration, threshold-based, and decision-curve analyses were based on one complete run of stratified 10-fold cross-validation, so that each participant had exactly one recalibrated out-of-fold prediction. Performance estimates derived from this procedure therefore represent internal validation and not external validation.

Discrimination was assessed using the area under the receiver operating characteristic curve (AUROC) and the area under the precision-recall curve (AUPRC). Calibration was assessed using the Brier score, the scaled Brier score (one minus the ratio of the Brier score to that of a null model assigning the observed prevalence to all participants), reliability plots, calibration intercept, calibration slope, calibration-in-the-large, and the observed-to-expected (O/E) ratio. To provide interpretable operating characteristics, we also evaluated the upper quintile and upper decile of recalibrated out-of-fold predicted risk as fixed risk strata, and calculated the corresponding predicted-risk cutoff, screen-positive proportion, sensitivity, specificity, positive predictive value, and negative predictive value. Decision-curve analysis was additionally performed across a range of threshold probabilities.

For model interpretation, features were ranked according to the absolute magnitude of standardized logistic regression coefficients averaged across cross-validation folds. Because the dietary total scores and their component scores were entered jointly, individual dietary coefficients should not be interpreted as independent effects. To explore the contribution of periconceptional diet to early-pregnancy prediction, we performed sequential ablation of Healthy Eating Index and Alternative Healthy Eating Index components in Model 1 and re-evaluated model performance after removal of each component.

The same modeling framework was applied to the LGA90 and LGA97 sensitivity outcomes, with discrimination and calibration summarized using AUROC, AUPRC, Brier score, calibration intercept, and calibration slope; confidence intervals for discrimination were obtained as described above. As an additional model-form sensitivity analysis, we compared the primary-outcome discrimination results with hyperparameter-tuned random forest, XGBoost, and gradient boosting models, using AUROC and AUPRC from cross-validated out-of-fold predictions.

### 2.6 Statistical analysis

Because this secondary analysis used an existing cohort, no a priori sample-size calculation was performed. Following Riley et al. [32], we report the available sample size (N=6,371) and event counts (>4000 g: 505; >4500 g: 77) in relation to the number of candidate predictors. The four models included 42, 51, 56, and 65 candidate predictors (Table 2), corresponding to 7.8–12.0 events per candidate predictor for birthweight >4000 g and 1.2–1.8 events per candidate predictor for birthweight >4500 g. Because penalized estimation does not use all candidate predictors to the same extent, we additionally examined model complexity for the Combined model for birthweight >4500 g. We estimated the local approximate effective degrees of freedom of the penalized model and evaluated two reduced versions of the Combined model, retaining 13 and 10 predictors, respectively. In each reduced model, predictors were ranked and selected separately within each outer training fold using their absolute standardized mean difference (Cohen’s d). This analysis used 100 repetitions of stratified 10-fold cross-validation, with Platt recalibration fitted on inner 5-fold cross-validated predictions within each outer training fold. The reported local effective degrees of freedom and calibration measures were summarized from a separate 100-repetition run. Results for birthweight >4500 g were interpreted cautiously given the limited number of events.

Baseline characteristics were summarized as mean (standard deviation) or number (percentage), as appropriate. Continuous variables were compared using Student’s t-test or Mann–Whitney U test according to distributional assumptions, and categorical variables were compared using chi-square test or Fisher’s exact test, as appropriate. Effect sizes were reported alongside significance tests. To account for multiple comparisons, p-values were adjusted using the Benjamini–Hochberg false discovery rate procedure. All tests were two-sided, and statistical significance was defined as p<0.05 after adjustment where applicable (q<0.05). Detailed univariate testing results, including raw p-values, adjusted q-values, and effect sizes, are provided in the Supplementary Material. Participant flow was summarized using sequential, mutually exclusive exclusion steps. To characterize potential selection during cohort derivation, we compared the final analytic cohort with the two main sequential exclusion groups: pregnancies excluded because of missing periconceptional diet data and pregnancies excluded because of incomplete Visit 2 and/or Visit 3 fetal biometry data after diet data were available. Because these two exclusion mechanisms may reflect different sources of selection, each excluded group was compared separately with the included analytic cohort across maternal sociodemographic characteristics, early-pregnancy clinical characteristics, and available pregnancy outcomes. For a limited additional evaluation in an independent Dutch cohort from Máxima Medical Center (MMC) [33], we developed a reduced early-pregnancy model in nuMoM2b using six predictors available in both datasets: maternal age, pre-pregnancy BMI, and four race and ethnicity indicators. The model was applied to the MMC delivery records with unchanged coefficients and intercept; records with missing data were excluded. This limited evaluation assessed discrimination only. The MMC data included multiparous women and repeated deliveries, and absolute-risk calibration was not evaluated. Reporting was checked against the TRIPOD+AI 2024 guidance for clinical prediction model studies [34]; the completed checklist is provided in Supplementary Table S11. Analyses were performed in Python 3.8 using scikit-learn 1.3.2.

## 3. Results

### 3.1 Cohort characteristics

Among the 10,038 women enrolled in the nuMoM2b cohort, 6,371 nulliparous pregnancies were included after excluding those lacking consent (N=749), outcome data (N=755), periconceptional diet information (N=1,349), or Visit 2/3 ultrasound measurements (N=814), as shown in Supplementary Figure S4. Because periconceptional diet and ultrasound data at both subsequent visits were required for the primary analysis, we further compared the included cohort with pregnancies excluded because of missing diet data or incomplete Visit 2/3 ultrasound follow-up (Supplementary Table S6). Excluded pregnancies differed from included pregnancies in several maternal sociodemographic characteristics and pregnancy outcomes, indicating potential selection related to data completeness and follow-up. Birthweight >4000 g occurred in 505 neonates (7.9%), including 77 (1.2%) >4500 g. Preterm birth occurred in 880 pregnancies (13.8%), and 284 (4.5%) were complicated by gestational diabetes. Pregnancy characteristics are shown in Table 1. Compared with non-macrosomic pregnancies, those with macrosomia generally exhibited higher maternal anthropometry, greater gestational weight gain, and larger later-pregnancy fetal biometry. These differences were more pronounced for birthweight >4500 g (Supplementary Tables S1 and S2).

### 3.2 Model performance and comparison with fetal growth chart–based benchmarks

Receiver operating characteristic curves for the four logistic regression models are shown in Figure 1. For prediction of birthweight >4000 g, Model 3 and the Combined model showed the highest discrimination, with AUROCs of 0.733 (95% confidence interval 0.709–0.755) and 0.732 (0.707–0.754), respectively. Compared with Model 1 (0.628, 0.602–0.651), the paired AUROC difference was 0.105 (0.081–0.128) for Model 3 and 0.104 (0.081–0.129) for the Combined model. Model 2 (0.651, 0.625– 0.674) showed a smaller improvement over Model 1 (difference 0.023, 0.013–0.037), and the Combined model did not improve on Model 3 (difference −0.001, −0.004 to 0.002). The estimates for Model 1 were obtained in the common analytic cohort with complete follow-up to Visit 3. Paired AUROC differences for all model comparisons are provided in Supplementary Table S5.

**Figure 1.**
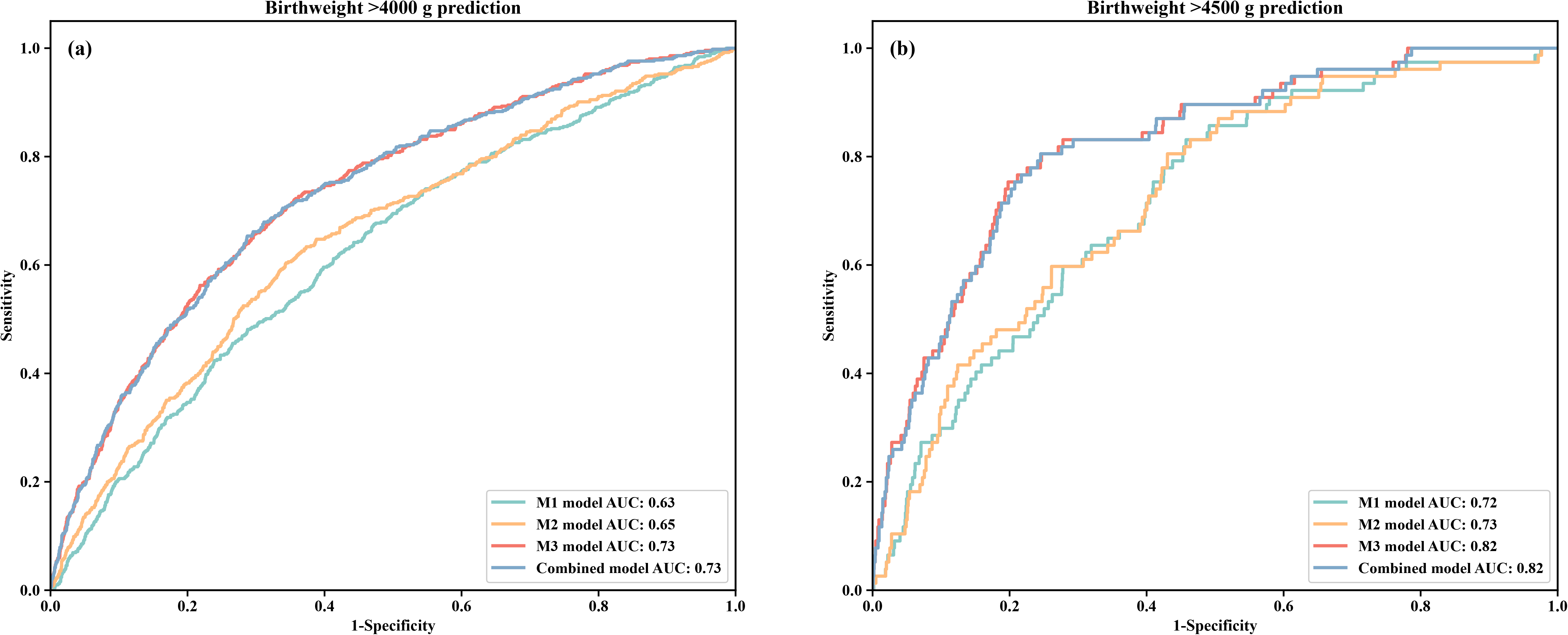
Receiver operating characteristic curves of the visit-updated logistic regression models for prediction of macrosomia. (a) Prediction of birthweight >4000 g. (b) Prediction of birthweight >4500 g.

WHO fetal growth chart–based percentile variables yielded lower discrimination than Model 3 and the Combined model. At Visit 2, the AUROC ranged from 0.54 to 0.59 for prediction of birthweight >4000 g. At Visit 3, performance improved to a range of 0.61–0.70, with the highest value observed for abdominal circumference percentile (0.70) and a value of 0.68 for estimated fetal weight percentile. However, Model 1 showed lower discrimination than the stronger Visit-3 WHO-derived percentiles. Full WHO chart–based results are summarized in Table 3.

**Table 3.** Area under the receiver operating characteristic curve of WHO fetal growth chart–based percentile variables for predicting macrosomia.

| WHO Characteristics | AUC | WHO Characteristics | AUC |
| --- | --- | --- | --- |
| For birthweight > 4000 g prediction |  | For birthweight > 4500 g prediction |  |
| WHO-BPD-2 | 0.57 | WHO-BPD-2 | 0.58 |
| WHO-AC-2 | 0.59 | WHO-AC-2 | 0.64 |
| WHO-HC-2 | 0.58 | WHO-HC-2 | 0.62 |
| WHO-FL-2 | 0.54 | WHO-FL-2 | 0.54 |
| WHO-EFW-2 | 0.57 | WHO-EFW-2 | 0.60 |
| WHO-BPD-3 | 0.65 | WHO-BPD-3 | 0.74 |
| WHO-AC-3 | 0.70 | WHO-AC-3 | 0.79 |
| WHO-HC-3 | 0.65 | WHO-HC-3 | 0.74 |
| WHO-FL-3 | 0.61 | WHO-FL-3 | 0.68 |
| WHO-EFW-3 | 0.68 | WHO-EFW-3 | 0.77 |

AUROC values are presented for descriptive comparison with the prediction models.
WHO: World Health Organization; BPD: biparietal diameter; AC: abdominal circumference; HC: head circumference; FL: femur length; EFW: estimated fetal weight. The suffixes 2 and 3 denote Visit 2 and Visit 3.

For prediction of birthweight >4500 g, Model 3 and the Combined model again showed the best discrimination, with AUROCs of 0.823 (0.775–0.864) and 0.821 (0.778–0.866), respectively, compared with 0.721 (0.670–0.770) for Model 1 and 0.727 (0.675–0.766) for Model 2. The paired AUROC difference relative to Model 1 was 0.102 (0.064–0.142) for Model 3 and 0.101 (0.061–0.139) for the Combined model, whereas Visit-2 information added little (Model 2 vs Model 1: 0.006, −0.006 to 0.019; Combined model vs Model 3: −0.002, −0.004 to 0.001). WHO chart–based percentile variables yielded AUROCs of 0.54–0.64 at Visit 2 and 0.68–0.79 at Visit 3. Among the WHO-derived measures, estimated fetal weight percentile was the most clinically interpretable single comparator (0.77 at Visit 3), whereas abdominal circumference percentile provided the highest Visit-3 AUROC (0.79). As for birthweight >4000 g, Model 1 showed lower discrimination than these Visit-3 percentiles, whereas the point estimates for Model 3 and the Combined model were higher. A supplementary comparison using INTERGROWTH-21st fetal ultrasound percentiles is provided in Supplementary Table S10. The INTERGROWTH-21st comparators showed a similar descriptive pattern to the WHO chart–based comparators, with stronger discrimination for Visit 3 than Visit 2 ultrasound percentiles. For birthweight >4000 g, Visit-3 abdominal circumference and estimated fetal weight percentiles achieved AUROCs of 0.71 and 0.69, respectively. For birthweight >4500 g, the corresponding AUROCs were 0.80 and 0.79. These results were closely aligned with the WHO-based comparator pattern, in which abdominal circumference and estimated fetal weight–related percentiles were among the strongest single ultrasound measures.

Additional performance metrics for the final models are provided in Supplementary Table S5 and Supplementary Figure S2. For the Combined model, the AUPRC was 0.209 (95% CI 0.181–0.240) for birthweight >4000 g and 0.066 (0.042–0.107) for birthweight >4500 g (Supplementary Figure S1). After Platt recalibration, calibration slopes ranged from 0.992 to 1.017, calibration-in-the-large from −0.018 to −0.001, and O/E ratios from 0.983 to 0.999 across the four models for both outcomes. The Combined model had a Brier score of 0.0686 (scaled Brier score, 6.0%) for birthweight >4000 g and 0.0116 (scaled Brier score, 2.9%) for birthweight >4500 g; these values indicate modest reductions in prediction error relative to the prevalence-only model. Supplementary Figure S2 shows the corresponding calibration plots.

Threshold-based operating characteristics are shown in Supplementary Table S7. For birthweight >4000 g, the Combined model classified 1,275 pregnancies as screen-positive at the upper-quintile threshold and captured 239 of 505 cases, with a sensitivity of 47.3%, specificity of 82.3%, positive predictive value (PPV) of 18.7%, and negative predictive value (NPV) of 94.8%. At the upper-decile threshold, it captured 147 of 505 cases, with a sensitivity of 29.1%, specificity of 91.6%, PPV of 23.0%, and NPV of 93.8%.

For birthweight >4500 g, Model 3 and the Combined model had similar operating characteristics. At the upper-quintile threshold, Model 3 captured 56 of 77 cases, with a sensitivity of 72.7%, specificity of 80.6%, PPV of 4.4%, and NPV of 99.6%; the Combined model captured 54 of 77 cases, with a sensitivity of 70.1%, specificity of 80.6%, PPV of 4.2%, and NPV of 99.5%. At the upper-decile threshold, Model 3 captured 36 of 77 cases, with a sensitivity of 46.8%, specificity of 90.4%, PPV of 5.6%, and NPV of 99.3%; the Combined model captured 35 of 77 cases, with a sensitivity of 45.5%, specificity of 90.4%, PPV of 5.5%, and NPV of 99.3%.

Decision-curve analysis for the two primary macrosomia outcomes is shown in Supplementary Figure S5. The decision curves summarize net benefit across threshold-probability ranges using recalibrated out-of-fold predicted probabilities and are intended to illustrate model behavior rather than to establish clinical utility or define recommended intervention thresholds.

In the model-complexity analysis for birthweight >4500 g (Supplementary Table S13), the local approximate effective degrees of freedom of the penalized Combined model was 24.2, compared with 65 candidate predictors. The 13-predictor model had local approximate effective degrees of freedom of 3.7 and an AUROC of 0.813 (95% CI 0.764–0.859). The corresponding values for the 10-predictor model were 3.0 and 0.803 (95% CI 0.752–0.851). The full model had an AUROC of 0.821 (95% CI 0.778–0.866) within the same framework. The paired AUROC differences from the full model were −0.008 (95% CI −0.037 to 0.018) for the 13-predictor model and −0.018 (95% CI −0.050 to 0.014) for the 10-predictor model. Their calibration slopes after Platt recalibration were 1.056 and 1.010, respectively.

Sensitivity analyses using gestational-age-adjusted LGA outcomes supported the same staged prediction pattern. LGA90 occurred in 1,319 pregnancies (20.7%) and LGA97 in 470 pregnancies (7.4%). For LGA90, AUROC increased from 0.620 (95% CI 0.604–0.637) in Model 1 to 0.731 (0.716–0.745) in Model 3. For LGA97, AUROC increased from 0.651 (0.626–0.675) to 0.758 (0.734–0.780). AUPRC also improved after incorporation of later ultrasound information. Full discrimination and calibration results for the LGA sensitivity outcomes are provided in Supplementary Table S4, with calibration plots shown in Supplementary Figure S3.

A separate model-form comparison using random forest, XGBoost, and gradient boosting is reported in Supplementary Table S8. These non-linear classifiers followed the same visit-updated performance pattern and did not show a consistent advantage over logistic regression.

In the limited evaluation in the MMC cohort (12,043 delivery records; 1,142 [9.5%] with birthweight >4000 g and 134 [1.1%] with birthweight >4500 g), the reduced six-predictor early-pregnancy model applied with the nuMoM2b coefficients yielded AUROCs of 0.597 (0.580–0.613) and 0.636 (0.589–0.681), respectively. In internal validation in nuMoM2b, the same reduced model achieved AUROCs of 0.603 (0.577–0.627) and 0.615 (0.556–0.678), which were lower than those of the full Model 1 (Supplementary Table S12).

### 3.3 Feature contributions of the combined model

The 20 predictors with the largest absolute standardized logistic regression coefficients in the Combined model are shown in Figures 2 and 3 for birthweight >4000 g and >4500 g, respectively. Overall, the most influential predictors were concentrated in maternal anthropometry, gestational weight gain, and second-and third-trimester fetal biometry. These results were consistent with the univariate analyses and support the incremental value of incorporating later ultrasound measurements into the visit-updated prediction framework. Coefficients are presented as mean and standard deviation across cross-validation folds.

**Figure 2.**
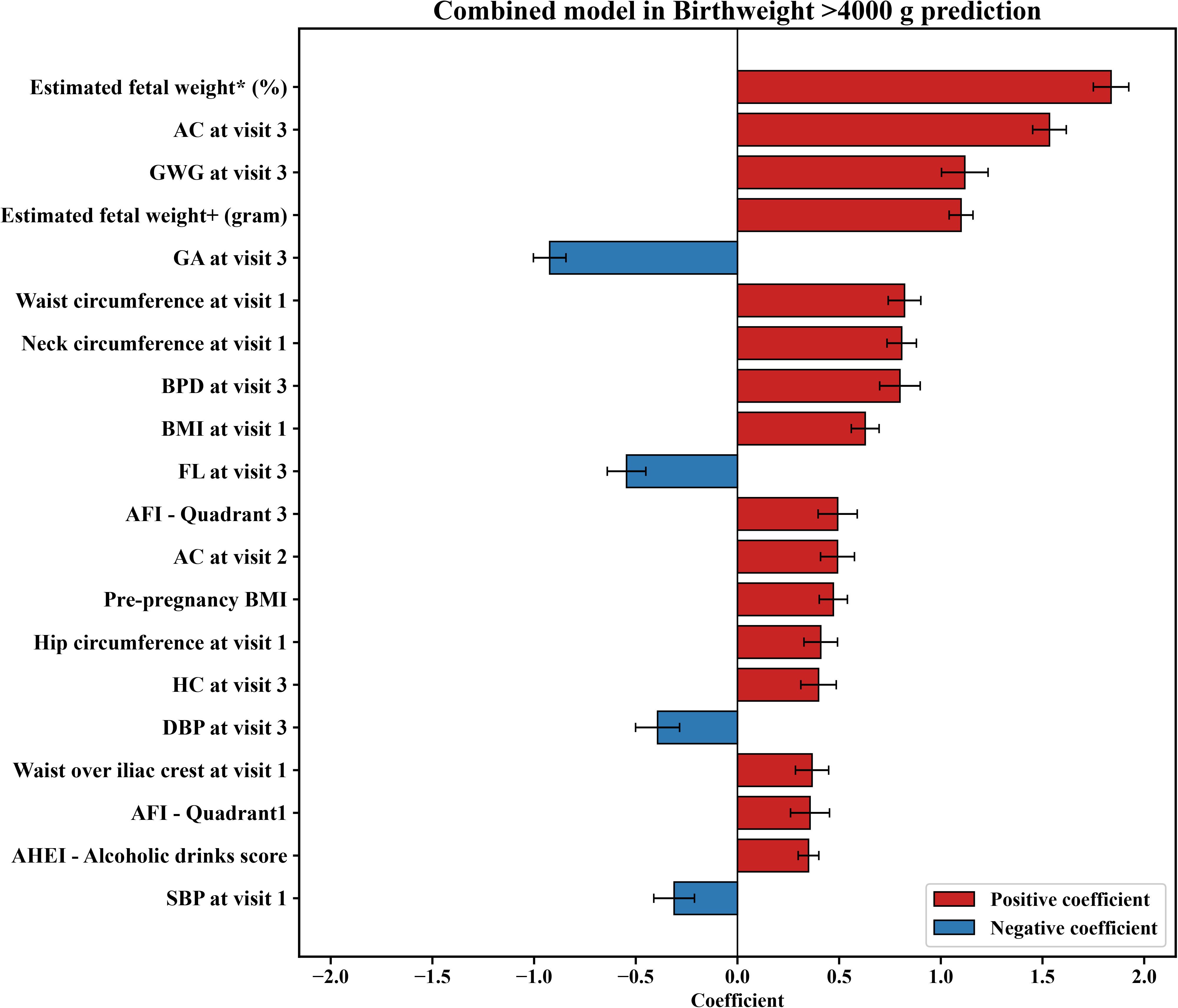
Twenty predictors with the largest absolute standardized logistic regression coefficients in the Combined model for prediction of birthweight >4000g. Bars extending to the right indicate positive coefficients and bars extending to the left indicate negative coefficients. Error bars represent standard deviation across cross-validation folds.

**Figure 3.**
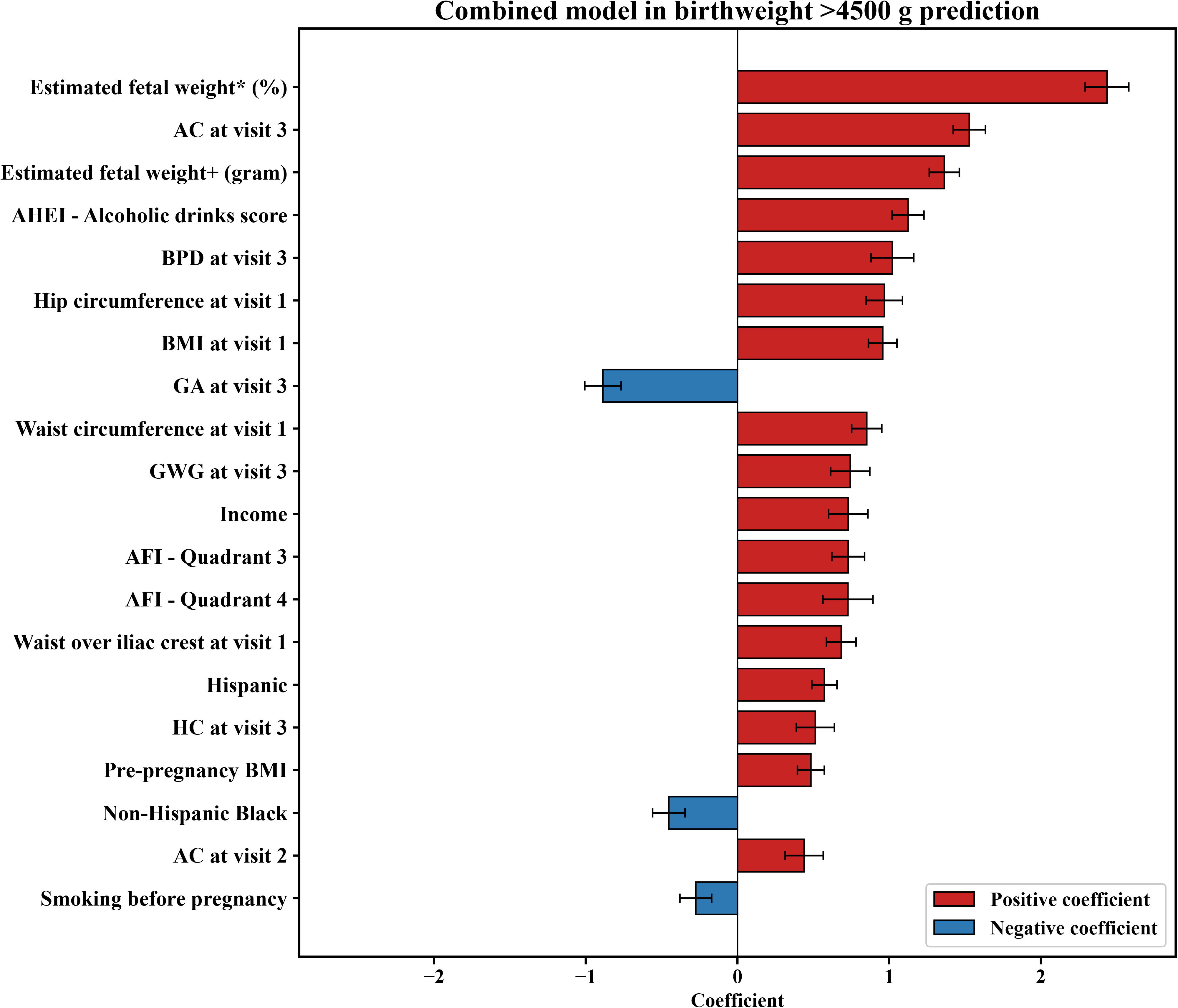
Twenty predictors with the largest absolute standardized logistic regression coefficients in the Combined model for prediction of birthweight >4500g. Bars extending to the right indicate positive coefficients and bars extending to the left indicate negative coefficients. Error bars represent standard deviation across cross-validation folds.

### 3.4 Periconceptional diet component analysis

The distribution of total diet quality scores is shown in Figure 4. Pregnancies resulting in macrosomia generally had slightly higher overall periconceptional diet quality scores; however, component-level comparisons indicated that most effect sizes were small (Supplementary Tables S1–S2). Exploratory analyses of changes in Model 1 discrimination after removal of dietary components are presented in Supplementary Figures S6 and S7.

**Figure 4.**
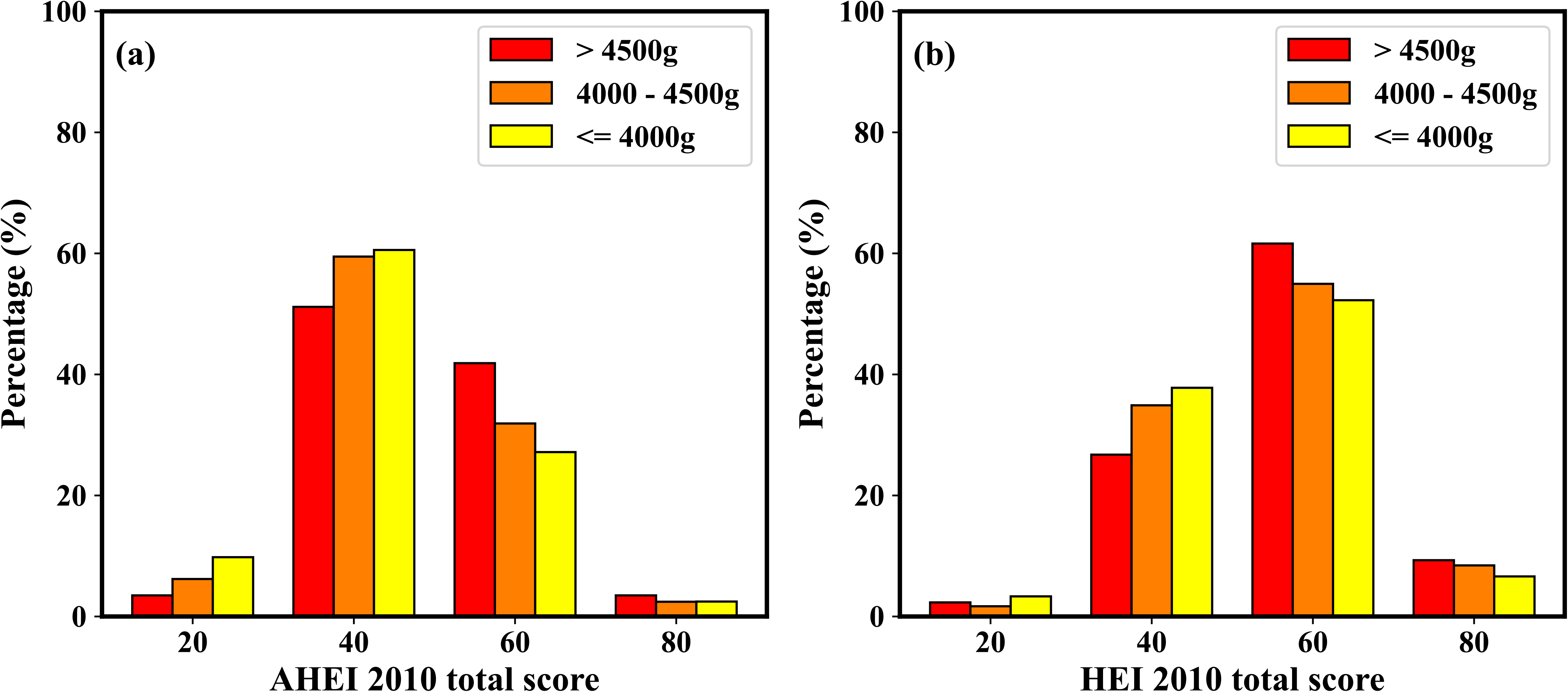
Distribution of total periconceptional diet quality scores according to birthweight category. (a) Alternative Healthy Eating Index-2010 total score. (b) Healthy Eating Index-2010 total score. Scores are shown for pregnancies with birthweight >4500 g, birthweight >4000 to ≤4500 g, and birthweight ≤4000 g.

## 4. Discussion

### 4.1 Principal findings

In this secondary analysis of a large prospective cohort of nulliparous singleton pregnancies in the United States, we developed and internally validated visit-updated multivariable logistic regression models for predicting macrosomia. Three main findings emerged. First, routinely available early-pregnancy maternal characteristics and anthropometric measurements provided moderate risk stratification for birthweight >4000 g and >4500 g. Second, model performance improved when second-and third-trimester ultrasound information was added, with the best discrimination observed for Model 3 and the Combined model. Third, exploratory diet-component analyses suggested that most individual dietary components contributed limited incremental predictive information.

Although the early-pregnancy model did not reach the discrimination of the stronger third-trimester WHO chart–based ultrasound percentiles, it relies on information that is available several months earlier in pregnancy. Once Visit-3 ultrasound information was incorporated, the visit-updated models showed higher point estimates than all WHO chart–based comparators, and most of the incremental discrimination was attributable to Visit-3 rather than Visit-2 information. Taken together, these findings support the potential role of early maternal data for initial risk stratification and of later ultrasound data for refinement of risk estimation across gestation.

Sensitivity analyses using LGA90 and LGA97 showed a similar visit-updated pattern, even though LGA and fixed birthweight thresholds represent related but distinct definitions of fetal overgrowth. Discrimination improved after incorporation of Visit-3 ultrasound information for both LGA outcomes, suggesting that the incremental value of later pregnancy information was not limited to fixed birthweight-threshold outcomes. The present analysis focused on birthweight-defined fetal overgrowth rather than downstream delivery complications such as postpartum hemorrhage, shoulder dystocia, or birth trauma, which may also depend on delivery management and other clinical factors beyond fetal size alone.

### 4.2 Comparison with previous studies

Our findings are consistent with previous studies showing that third-trimester ultrasound generally provides better discrimination for macrosomia than earlier ultrasound measurements [13,14]. This is clinically plausible, because fetal overgrowth becomes more apparent with advancing gestation and later biometric measurements better capture cumulative growth patterns. At the same time, our results highlight the limitations of relying only on later-pregnancy ultrasound, because such an approach leaves less opportunity for earlier risk stratification and preventive counseling.

Compared with prior prediction studies, our study adds to the literature in three ways. First, it focuses specifically on nulliparous pregnancies, in which prediction is more challenging because previous obstetric history is unavailable [35]. Second, it evaluates a visit-updated framework that reflects how information becomes available during routine antenatal care, rather than relying solely on near-term ultrasound [13–18]. Third, it benchmarks model performance against WHO fetal growth chart–based ultrasound percentiles, thereby providing a clinically interpretable reference for comparison [26,27]. The present study therefore provides a clinically oriented prediction framework developed and internally validated in a large prospective cohort, while further external validation is needed before routine clinical implementation.

Our dietary findings should be interpreted in the context of previous observational work. Existing evidence on the relation between maternal diet quality and macrosomia has been mixed [36–39]. In the nuMoM2b cohort, Yee et al. previously reported that higher pre-pregnancy diet quality was associated with increased odds of macrosomia >4000 g [40]. Our results are broadly consistent with this observation at the overall diet-quality level, but the component-level analyses suggested that most individual dietary components contributed little incremental predictive information when considered separately.

### 4.3 Clinical implications

The clinical value of early macrosomia prediction lies less in replacing ultrasound and more in enabling earlier risk stratification. In routine care, pregnancies identified as being at increased risk could be considered for closer monitoring of gestational weight gain, more individualized nutritional counseling, and greater attention to fetal growth across follow-up visits [9,41–45]. This may be particularly relevant in nulliparous pregnancies, where clinicians cannot rely on prior delivery history to guide risk assessment [35].

Our findings also suggest that a staged strategy may be more realistic than a single-time-point prediction approach. Early-pregnancy maternal data may be useful for initial screening, whereas later ultrasound measurements can refine risk estimates as pregnancy progresses. This interpretation is more clinically appropriate than positioning the model as a replacement for standard sonographic assessment. Rather, the proposed framework may complement existing care by identifying higher-risk pregnancies earlier than ultrasound-only approaches.

The threshold-based analyses translate the discrimination results into operating characteristics at fixed screening fractions. For birthweight >4000 g, 23.0% of pregnancies in the highest decile of Combined-model predicted risk had the outcome, compared with an overall prevalence of 7.9%. This indicates that the upper-decile group had a higher observed event rate than the cohort average, although sensitivity at this threshold was 29.1%. For birthweight >4500 g, the highest-risk strata also showed higher observed event rates than the overall prevalence of 1.2%, but the absolute positive predictive value remained modest, reaching 5.6% in the upper decile. These results are therefore best interpreted as describing screening yield at fixed risk strata, rather than as defining thresholds for clinical intervention. Because no specific intervention or harm–benefit trade-off was defined, neither these analyses nor the decision-curve analysis establish clinical utility.

### 4.4 Strengths and limitations

This study has several strengths. It was based on a large prospective multicenter cohort with standardized antenatal data collection [19]. The focus on nulliparous singleton pregnancies addresses a clinically important population in which macrosomia risk prediction is often more difficult. In addition, the visit-updated modeling strategy reflects the sequential availability of clinical information in routine care. Finally, we reported not only discrimination but also precision–recall performance and calibration, which strengthens the transparency of model evaluation.

Several limitations should also be acknowledged. First, this was a secondary analysis and performance estimates for the four full models were derived from internal validation only; external validation in independent populations is required before clinical implementation. In a limited evaluation in an independent Dutch cohort, a reduced early-pregnancy model restricted to six predictors available in both datasets showed discrimination similar to that observed for the same reduced model in nuMoM2b (Supplementary Table S12). This analysis did not include periconceptional diet, most Visit-1 anthropometric measurements, or serial ultrasound measurements, included multiparous women, and assessed discrimination only; it therefore does not constitute external validation of the full visit-updated models. Second, the cohort consisted of nulliparous singleton pregnancies from the United States, which may limit generalizability to other populations and care settings. Third, ultrasound data near delivery were not available, which restricts comparison with models based on near-term fetal biometry and may underestimate the performance achievable with later gestational data. In addition, because estimated fetal weight was derived using the Hadlock formula, possible underestimation of fetal weight at the upper extremes should be considered when interpreting EFW-based predictors and chart-based comparators. Relatedly, although we added LGA90 and LGA97 sensitivity analyses to account for gestational age at delivery, absolute macrosomia and LGA should not be interpreted interchangeably. Future studies should determine which fetal overgrowth definition best predicts delivery and neonatal complications. Fourth, periconceptional dietary intake was assessed using the Block 2005 Food Frequency Questionnaire, which relied on self-reported recall of usual intake during the 3 months around conception and was therefore subject to recall bias and exposure misclassification. Dietary intake was not assessed longitudinally across pregnancy, limiting inference about changes in diet over time. Restricting the cohort to pregnancies with periconceptional diet and serial Visit 2/Visit 3 ultrasound data may also have introduced selection bias. Although this restriction was necessary for evaluating visit-updated models incorporating diet and antenatal ultrasound information, the included-versus-excluded comparison suggested that data completeness and follow-up were not entirely random. In particular, Model 1 was evaluated only among pregnancies with periconceptional dietary data and subsequent Visit-2 and Visit-3 ultrasound data, so its performance may not reflect that in all pregnancies eligible at Visit 1. In addition, prediction of birthweight >4500 g was based on only 77 events; although the model-complexity analysis suggested that most of the discrimination was retained with substantially fewer predictors, estimates for this outcome might be imprecise and should be interpreted cautiously. Fifth, although the WHO fetal growth chart provided an interpretable benchmark, direct comparison with our internally derived models should be made cautiously because the WHO chart was developed from a different reference population and was not calibrated to the same dataset.

## 5. Conclusion

In the selected analytic cohort of nulliparous singleton pregnancies, routinely available early-pregnancy maternal characteristics and anthropometry provided moderate risk stratification for macrosomia, while later ultrasound measurements improved performance further. The proposed visit-updated framework may support earlier identification of pregnancies at increased risk, but external validation is needed before clinical use.

## Declarations

## Ethics Statement

This study analyzed de-identified data from the nuMoM2b dataset. The Eunice Kennedy Shriver National Institute of Child Health and Human Development review board (approval date: December 15, 2021, request ID: 11961) and Eindhoven University of Technology ethical review board (approval date: November 1, 2021, reference number: ERB2021BME4) and the Medical Ethics Review Committee of Máxima Medical Center approved the study protocol [33]. Our study complied with all relevant ethical regulations, including the Declaration of Helsinki and the Good Clinical Practice Guidelines.

## Data Availability Statement

The data that support the findings of this study are available from the Eunice Kennedy Shriver National Institute of Child Health and Human Development Data and Specimen Hub upon application and approval. Restrictions apply to the availability of these data. The MMC data are not publicly available because of privacy regulations; further information is available from the corresponding author upon reasonable request.

## Consent to participate

Informed consent was obtained from all subjects involved in the original nuMoM2b study.

## Author Contributions

**Yanqi Wu** contributed to the conceptualization, study design, data interpretation, statistical analysis, visualization, and drafting of the manuscript.

**Sima Asvadi** contributed to the conceptualization, study design, data interpretation, and critical revision of the manuscript.

**Myrthe van der Ven** contributed to the methodology, supervision, data interpretation, and critical revision of the manuscript.

**Elisabetta Peri** contributed to the methodology, supervision, statistical analysis, and critical revision of the manuscript.

**M. Beatrijs van der Hout-van der Jagt** contributed to data curation, statistical analysis, and critical revision of the manuscript.

**S. Guid Oei** contributed to the methodology, supervision, and critical revision of the manuscript.

**Massimo Mischi** was involved in project administration, supervision, and manuscript editing.

**Xi Long** contributed to project administration, supervision, investigation, conceptualization, study design, and critical revision of the manuscript. All authors read and approved the final manuscript.

