## Supplementary V1 for "Development and internal validation of early- and mid-pregnancy prediction models for macrosomia in nulliparous singleton pregnancies"

### Supplemental Materials

### Supplementary Methods: Derivation of WHO fetal growth chart–based percentile variables

For each pregnancy with an available ultrasound examination at Visit 2 or Visit 3, gestational age at scan and one fetal biometric measurement were mapped to the World Health Organization fetal growth chart to obtain a gestational-age-specific percentile. The biometric inputs included head circumference, biparietal diameter, abdominal circumference, femur length, and estimated fetal weight. Estimated fetal weight was first calculated from ultrasound biometry using the Hadlock formula and then converted into a WHO chart–based percentile. Percentiles were computed separately for Visit 2 and Visit 3. Accordingly, five WHO-derived percentile variables were generated at Visit 2 and five additional variables were generated at Visit 3. Variable names followed the format WHO-[biometric variable]-[visit], where the suffix "2" denotes Visit 2 and the suffix "3" denotes Visit 3. For example, WHO-EFW-2 represents the percentile derived from estimated fetal weight and gestational age at Visit 2, whereas WHO-AC-3 represents the percentile derived from abdominal circumference and gestational age at Visit 3.

All WHO-derived percentile variables were evaluated as ultrasound-based benchmark predictors in the comparison with the logistic regression models. Their discriminative performance was summarized as area under the receiver operating characteristic curve values in Table 3 of the main manuscript. These values are presented for descriptive comparison with the prediction models; no statistical tests were performed between the chart-based comparators and the prediction models. The estimated fetal weight percentile was additionally emphasized in the narrative interpretation because it most closely reflects routine clinical assessment of fetal overgrowth.

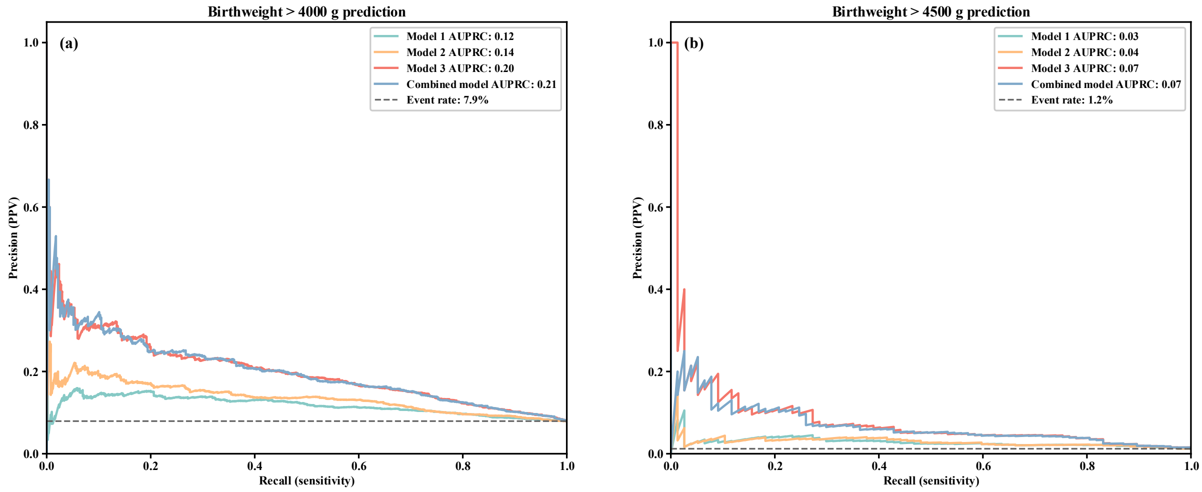


### Figure S1. Precision–recall (PR) curves for macrosomia prediction.

(a) >4000 g (prevalence 8%). (b) >4500 g (prevalence 1%). Curves were computed from participant-level out-of-fold predictions, averaged for each participant across the repeated runs of stratified 10-fold cross-validation. Models: Model 1 (Visit 1), Model 2 (Model 1 + Visit 2), Model 3 (Model 1 + Visit 3), and the Combined model (Visits 1–3); the predictors in each model are listed in Table 2 of the main manuscript. AUPRC (area under the precision–recall curve) values by model: >4000 g—M1 0.12, M2 0.14, M3 0.20, Combined 0.21; >4500 g—M1 0.03, M2 0.04, M3 0.07, Combined 0.07.

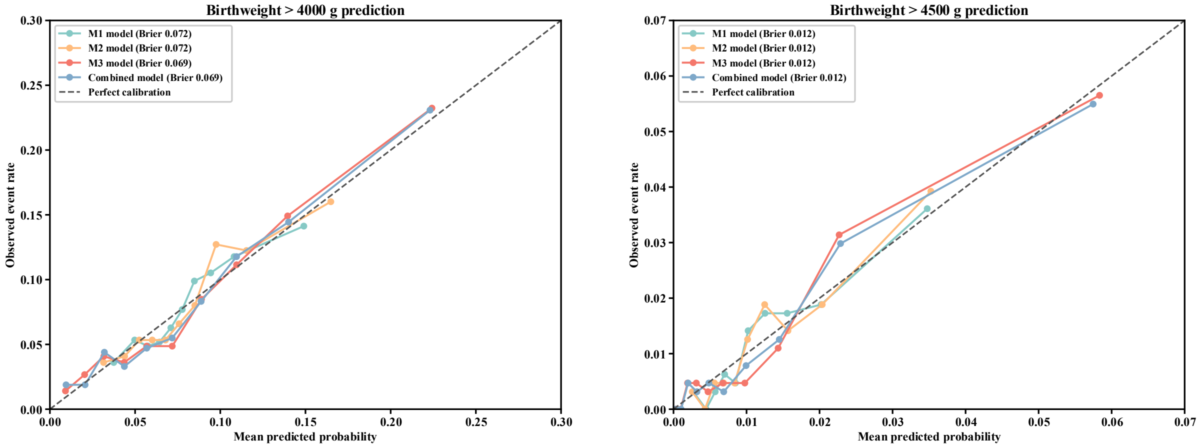


### Figure S2. Calibration plots for macrosomia prediction.

(a) >4000 g. (b) >4500 g. Calibration plots are based on Platt-recalibrated out-of-fold predicted probabilities from one complete run of stratified 10-fold cross-validation, in which the Platt calibrator was fitted within the training folds only; each participant contributes one prediction. Models: Model 1 (Visit 1), Model 2 (Model 1 + Visit 2), Model 3 (Model 1 + Visit 3), and the Combined model (Visits 1–3). Brier scores (a): M1 0.0721, M2 0.0715, M3 0.0686, Combined 0.0686. Brier scores (b): M1 0.0119, M2 0.0119, M3 0.0116, Combined 0.0116.

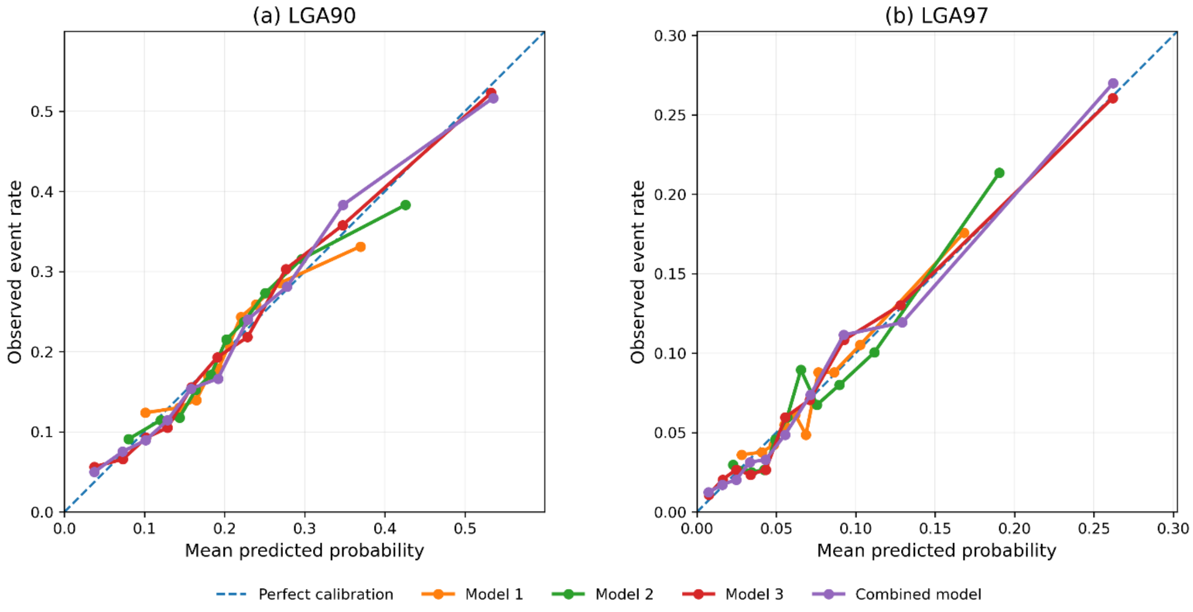


### Figure S3. Calibration plots for LGA sensitivity outcomes.

Calibration plots were generated from Platt-recalibrated out-of-fold predicted probabilities from one complete run of stratified 10-fold cross-validation, in which the Platt calibrator was fitted within the training folds only. Each point represents one predicted-risk block, plotted as the mean predicted probability against the observed event rate. The dashed diagonal line indicates perfect calibration. Panels show sensitivity outcomes defined as birthweight above the INTERGROWTH-21st gestational-age- and sex-specific (a) 90th percentile and (b) 97th percentile. Model 1, Model 2, Model 3, and the Combined model are shown separately.

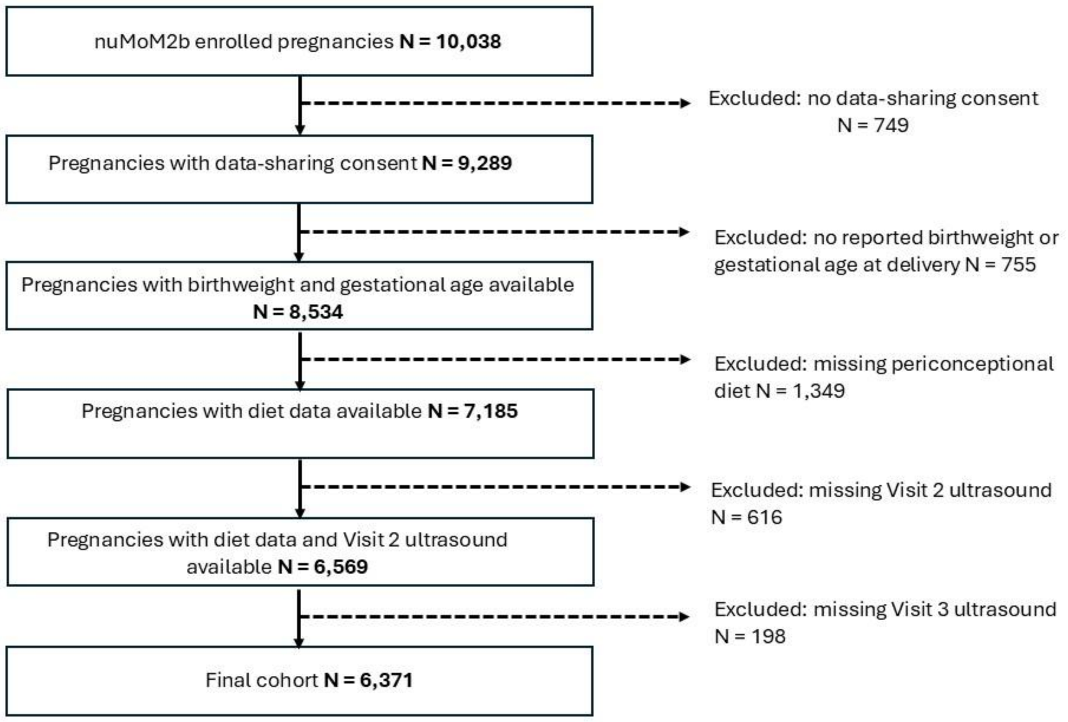


**Supplementary Figure S4. Participant flow for derivation of the analytic cohort.**

Note: Exclusions are shown sequentially and are therefore mutually exclusive. Among 10,038 women enrolled in the nuMoM2b cohort, 749 were excluded because data-sharing consent was unavailable. Among the remaining 9,289 pregnancies, 755 were excluded because birthweight or gestational age at delivery was not reported, leaving 8,534 pregnancies with outcome information. Of these, 1,349 were excluded because of missing periconceptional diet data. Among the remaining 7,185 pregnancies, 616 lacked Visit 2 ultrasound data, including 523 missing Visit 2 only and 93 missing both Visit 2 and Visit 3. Among the 6,569 pregnancies with Visit 2 ultrasound available, 198 additionally lacked Visit 3 ultrasound data. The final analytic cohort included 6,371 pregnancies.

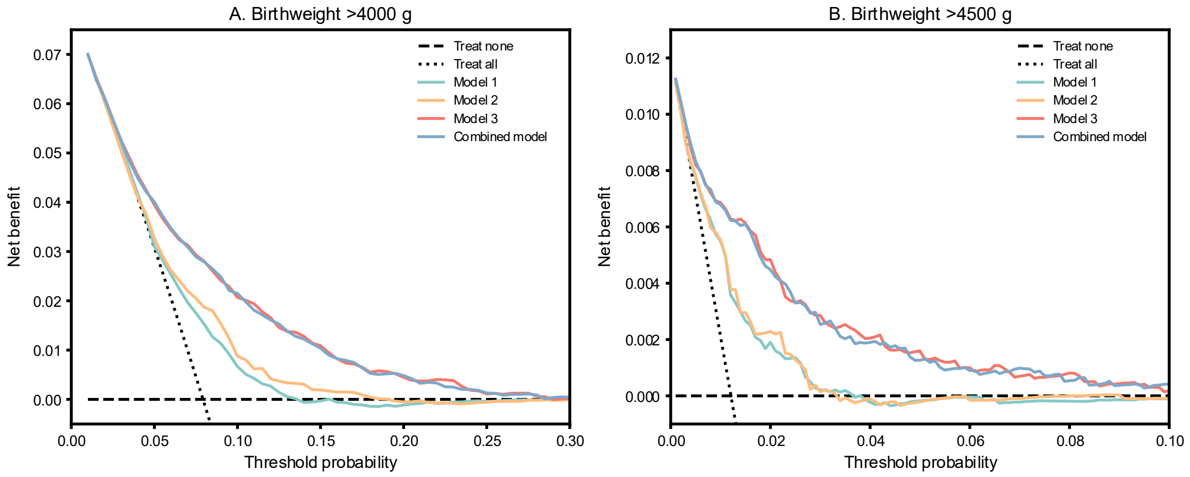

#### Supplementary Figure S5. Decision curve analysis for primary macrosomia outcomes.

Decision curve analysis was performed using Platt-recalibrated out-of-fold predicted probabilities from one complete run of stratified 10-fold cross-validation. Net benefit was calculated across threshold-probability ranges and compared with treat-all and treat-none reference strategies. Panels show decision curves for (a) birthweight >4000 g and (b) birthweight >4500 g. Because birthweight >4500 g was rare, a lower threshold-probability range was used for this outcome. The curves are intended to illustrate model behavior rather than to establish clinical utility or define recommended intervention thresholds, because no specific clinical intervention or harm–benefit trade-off was defined.

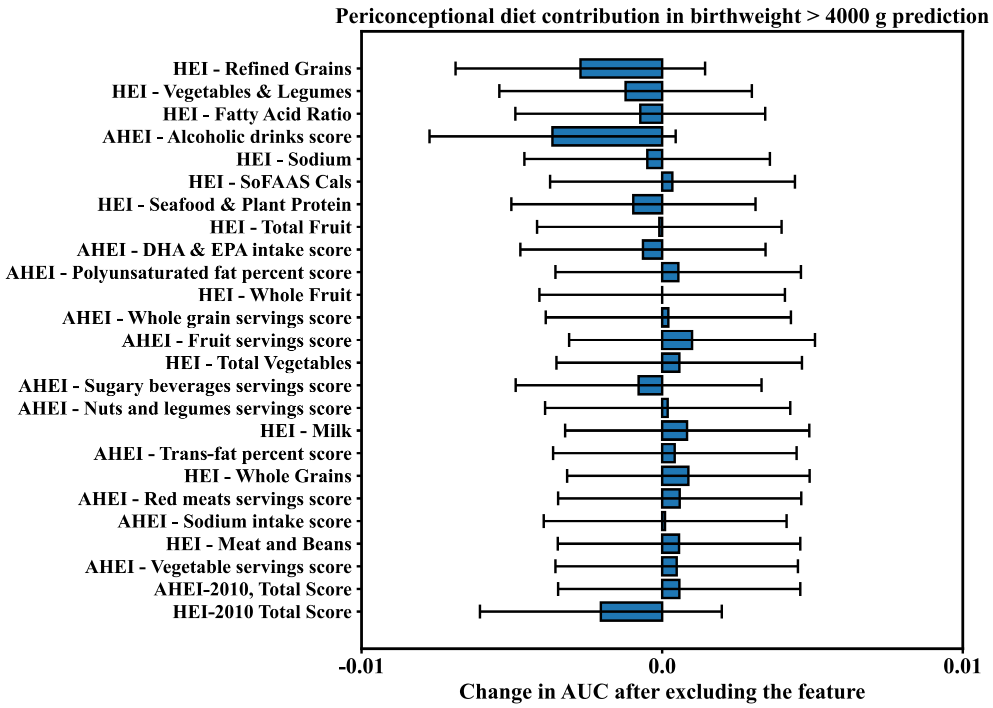


#### Supplementary Figure S6. Change in area under the receiver operating characteristic curve after sequential removal of individual periconceptional diet components from Model 1 for prediction of birthweight >4000 g.

Values are presented as mean and standard deviation across cross-validation runs, with standard deviation shown as error bars. This ablation analysis was exploratory.

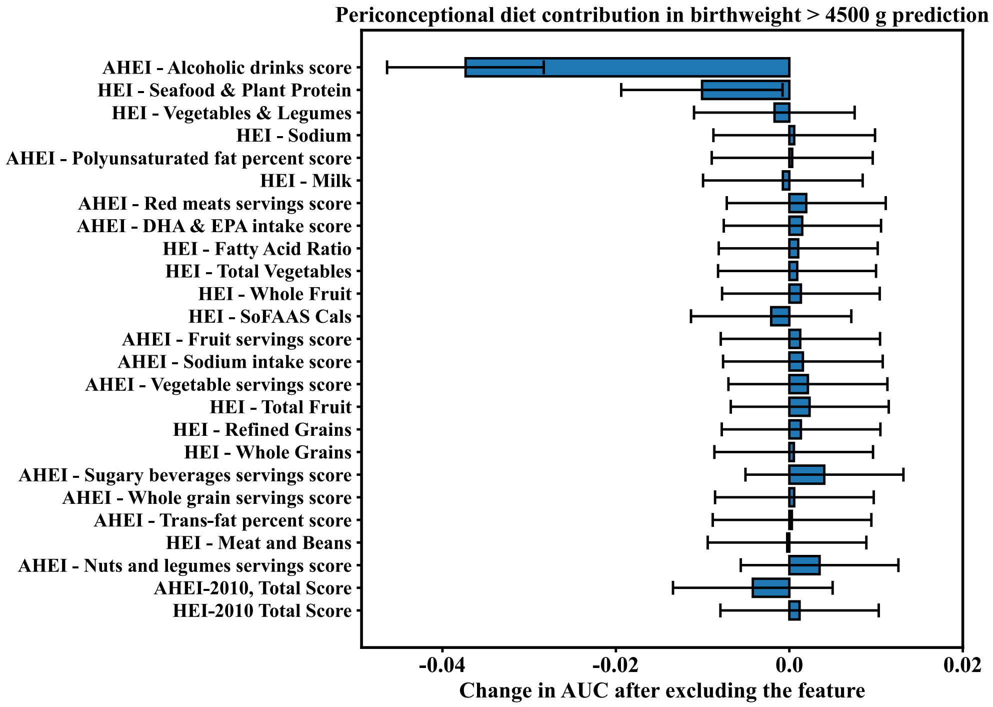


**Supplementary Figure S7. Change in area under the receiver operating characteristic curve after sequential removal of individual periconceptional diet components from Model 1 for prediction of birthweight >4500 g.**

Values are presented as mean and standard deviation across cross-validation runs, with standard deviation shown as error bars. This ablation analysis was exploratory. For Figures S6 and S7, the AHEI-2010 alcoholic drinks component uses a non-monotonic score (for women, 10 points for 0.5–1.5 drinks/day, 2.5 for no intake, and 0 for ≥2.5 drinks/day) [24]. Component-ablation results describe predictive contributions within the dietary predictor set and do not estimate independent exposure effects.

### Table S1. Univariate associations for macrosomia >4000 g.

For each predictor: test used (t-test with Welch as needed, Mann–Whitney U, or $\chi^2$/Fisher), effect size (Cramér's V/Cliff's delta/odds ratio), raw p, and BH-FDR q (q<0.05, two-sided α=0.05). Variable names/units follow Table 1.

| Variable | Type | Test | P value | Effect size | Effect type | Q value | Significant |
|---|---|---|---|---|---|---|---|
| Age | numeric | Mann–Whitney U | <0.001 | -0.105 | Cliff's delta | <0.001 | TRUE |
| Non-Hispanic white | categorical | Chi-square | <0.001 | 0.045 | Cramér's V | <0.001 | TRUE |
| Non-Hispanic black | categorical | Chi-square | <0.05 | 0.0322 | Cramér's V | <0.05 | TRUE |

| Variable | Type | Test | P value | Effect size | Effect type | Q value | Significant |
|---|---|---|---|---|---|---|---|
| Hispanic | categorical | Chi-square | 0.25 | 0.014 | Cramér's V | 0.30 | FALSE |
| Asian/Other race or ethnicity | categorical | Chi-square | 0.06 | 0.0235 | Cramér's V | 0.08 | FALSE |
| Income | numeric | Mann–Whitney U | <0.001 | -0.105 | Cliff's delta | <0.001 | TRUE |
| Smoke | categorical | Chi-square | 0.75 | 0.004 | Cramér's V | 0.77 | FALSE |
| Fetal sex | categorical | Chi-square | <0.001 | 0.089 | Cramér's V | <0.001 | TRUE |
| Pre-pregnancy BMI | numeric | Mann–Whitney U | <0.001 | -0.189 | Cliff's delta | <0.001 | TRUE |
| Preexisting diabetes | categorical | Chi-square | 0.42 | 0.010 | Cramér's V | 0.46 | FALSE |
| HEI - Total Vegetables | numeric | Mann–Whitney U | <0.05 | -0.065 | Cliff's delta | <0.05 | TRUE |
| HEI - Vegetables & Legumes | numeric | Mann–Whitney U | 0.26 | -0.028 | Cliff's delta | 0.31 | FALSE |
| HEI - Total Fruit | numeric | Mann–Whitney U | <0.05 | -0.080 | Cliff's delta | <0.05 | TRUE |
| HEI - Whole Fruit | numeric | Mann–Whitney U | <0.001 | -0.094 | Cliff's delta | <0.001 | TRUE |
| HEI - Whole Grains | numeric | Mann–Whitney U | <0.05 | -0.065 | Cliff's delta | <0.05 | TRUE |
| HEI - Milk | numeric | Mann–Whitney U | 0.10 | -0.043 | Cliff's delta | 0.13 | FALSE |
| HEI - Meat and Beans | numeric | Mann–Whitney U | 0.31 | -0.026 | Cliff's delta | 0.36 | FALSE |
| HEI - Seafood & Plant Protein | numeric | Mann–Whitney U | <0.05 | -0.059 | Cliff's delta | <0.05 | TRUE |
| HEI - Fatty Acid Ratio | numeric | Mann–Whitney U | 0.79 | 0.007 | Cliff's delta | 0.80 | FALSE |
| HEI - Sodium | numeric | Mann–Whitney U | 0.07 | 0.047 | Cliff's delta | 0.09 | FALSE |
| HEI - Refined Grains | numeric | Mann–Whitney U | <0.05 | -0.063 | Cliff's delta | <0.05 | TRUE |
| HEI - SoFAAS Cals | numeric | Mann–Whitney U | <0.05 | -0.073 | Cliff's delta | <0.05 | TRUE |

| Variable | Type | Test | P value | Effect size | Effect type | Q value | Significant |
|---|---|---|---|---|---|---|---|
| HEI - Total Score | numeric | Mann–Whitney U | <0.001 | -0.095 | Cliff's delta | <0.001 | TRUE |
| AHEI - Vegetable servings score | numeric | Mann–Whitney U | <0.05 | -0.065 | Cliff's delta | <0.05 | TRUE |
| AHEI - Fruit servings score | numeric | Mann–Whitney U | <0.001 | -0.096 | Cliff's delta | <0.001 | TRUE |
| AHEI - Whole grain servings score | numeric | Mann–Whitney U | <0.05 | -0.064 | Cliff's delta | <0.05 | TRUE |
| AHEI - Sugary beverages servings score | numeric | Mann–Whitney U | 0.15 | -0.036 | Cliff's delta | 0.18 | FALSE |
| AHEI - Nuts and legumes servings score | numeric | Mann–Whitney U | <0.05 | -0.056 | Cliff's delta | 0.05 | FALSE |
| AHEI - Red meats servings score | numeric | Mann–Whitney U | 0.57 | 0.015 | Cliff's delta | 0.60 | FALSE |
| AHEI - Trans-fat percent score | numeric | Mann–Whitney U | 0.23 | -0.032 | Cliff's delta | 0.27 | FALSE |
| AHEI - DHA & EPA intake score | numeric | Mann–Whitney U | 0.98 | 0.001 | Cliff's delta | 0.98 | FALSE |
| AHEI - Polyunsaturated fat percent score | numeric | Mann–Whitney U | 0.54 | -0.016 | Cliff's delta | 0.58 | FALSE |
| AHEI - Sodium intake score | numeric | Mann–Whitney U | 0.37 | 0.023 | Cliff's delta | 0.42 | FALSE |
| AHEI - Alcoholic drinks score | categorical | Chi-square | <0.001 | 0.057 | Cramér's V | <0.001 | TRUE |
| AHEI - Total Score | numeric | Mann–Whitney U | <0.001 | -0.093 | Cliff's delta | <0.05 | TRUE |
| BMI at Visit 1 | numeric | Mann–Whitney U | <0.001 | -0.207 | Cliff's delta | <0.001 | TRUE |
| waist circumference at Visit 1 | numeric | Mann–Whitney U | <0.001 | -0.098 | Cliff's delta | <0.001 | TRUE |
| waist over iliac crest at Visit 1 | numeric | Mann–Whitney U | <0.001 | -0.197 | Cliff's delta | <0.001 | TRUE |
| Hip circumference at Visit 1 | numeric | Mann–Whitney U | <0.001 | -0.202 | Cliff's delta | <0.001 | TRUE |
| Neck circumference at Visit 1 | numeric | Mann–Whitney U | <0.001 | -0.206 | Cliff's delta | <0.001 | TRUE |

| Variable | Type | Test | P value | Effect size | Effect type | Q value | Significant |
|---|---|---|---|---|---|---|---|
| SBP at Visit 1 | numeric | Mann–Whitney U | <0.001 | -0.233 | Cliff's delta | <0.001 | TRUE |
| DBP at Visit 1 | numeric | Mann–Whitney U | <0.05 | -0.053 | Cliff's delta | 0.06 | FALSE |
| GWG at Visit 1 | numeric | Mann–Whitney U | <0.05 | -0.060 | Cliff's delta | <0.05 | TRUE |
| GWG at Visit 2 | numeric | Mann–Whitney U | <0.001 | -0.147 | Cliff's delta | <0.001 | TRUE |
| SBP at Visit 2 | numeric | Mann–Whitney U | <0.05 | -0.070 | Cliff's delta | <0.05 | TRUE |
| DBP at Visit 2 | numeric | Mann–Whitney U | 0.06 | -0.049 | Cliff's delta | 0.08 | FALSE |
| GDM | categorical | Chi-square | 0.05 | 0.024 | Cramér's V | 0.07 | FALSE |
| GA at Visit 2 | numeric | Mann–Whitney U | <0.05 | -0.052 | Cliff's delta | 0.06 | FALSE |
| BPD at Visit 2 | numeric | Mann–Whitney U | <0.001 | -0.147 | Cliff's delta | <0.001 | TRUE |
| HC at Visit 2 | numeric | Mann–Whitney U | <0.001 | -0.149 | Cliff's delta | <0.001 | TRUE |
| AC at Visit 2 | numeric | Mann–Whitney U | <0.001 | -0.175 | Cliff's delta | <0.001 | TRUE |
| FL at Visit 2 | numeric | Mann–Whitney U | <0.001 | -0.109 | Cliff's delta | <0.001 | TRUE |
| EFW at Visit 2 | numeric | Mann–Whitney U | <0.001 | -0.152 | Cliff's delta | <0.001 | TRUE |
| GWG at Visit 3 | numeric | Mann–Whitney U | <0.001 | -0.183 | Cliff's delta | <0.001 | TRUE |
| SBP at Visit 3 | numeric | Mann–Whitney U | <0.05 | -0.053 | Cliff's delta | 0.06 | FALSE |
| DBP at Visit 3 | numeric | Mann–Whitney U | 0.44 | -0.020 | Cliff's delta | 0.48 | FALSE |
| GA at Visit 3 | numeric | Mann–Whitney U | <0.001 | -0.091 | Cliff's delta | 0.001 | TRUE |
| BPD at Visit 3 | numeric | Mann–Whitney U | <0.001 | -0.301 | Cliff's delta | <0.001 | TRUE |

| Variable | Type | Test | P value | Effect size | Effect type | Q value | Significant |
|---|---|---|---|---|---|---|---|
| HC at Visit 3 | numeric | Mann–Whitney U | <0.001 | -0.288 | Cliff's delta | <0.001 | TRUE |
| AC at Visit 3 | numeric | Mann–Whitney U | <0.001 | -0.362 | Cliff's delta | <0.001 | TRUE |
| FL at Visit 3 | numeric | Mann–Whitney U | <0.001 | -0.230 | Cliff's delta | <0.001 | TRUE |
| EFW at Visit 3 | numeric | Mann–Whitney U | <0.001 | -0.103 | Cliff's delta | <0.001 | TRUE |
| AFI - Quadrant 1 at Visit 3 | numeric | Mann–Whitney U | <0.001 | -0.131 | Cliff's delta | <0.001 | TRUE |
| AFI - Quadrant 2 at Visit 3 | numeric | Mann–Whitney U | <0.05 | -0.070 | Cliff's delta | <0.05 | TRUE |
| AFI - Quadrant 3 at Visit 3 | numeric | Mann–Whitney U | <0.001 | -0.335 | Cliff's delta | <0.001 | TRUE |
| AFI - Quadrant 4 at Visit 3 | numeric | Mann–Whitney U | <0.001 | -0.381 | Cliff's delta | <0.001 | TRUE |

**Table S2. Univariate associations for macrosomia >4500 g.**

For each predictor: test used (t-test with Welch as needed, Mann–Whitney U, or $\chi^2$/Fisher), effect size (Cramér's V/Cliff's delta/odds ratio), raw p, and BH-FDR q (q<0.05, two-sided α=0.05). Variable names/units follow Table 1.

| Variable | Type | Test | P value | Effect size | Effect type | Q value | Significant |
|---|---|---|---|---|---|---|---|
| Age | numeric | Mann–Whitney U | <0.001 | -0.202 | Cliff's delta | <0.05 | TRUE |
| Non-Hispanic white | categorical | Chi-square | 0.68 | 0.005 | Cramér's V | 0.74 | FALSE |
| Non-Hispanic black | categorical | Chi-square | 0.10 | 0.020 | Cramér's V | 0.19 | FALSE |
| Hispanic | categorical | Chi-square | 0.14 | 0.018 | Cramér's V | 0.23 | FALSE |
| Asian/Other race or ethnicity | categorical | Chi-square | 0.43 | 0.009 | Cramér's V | 0.51 | FALSE |
| Income | numeric | Mann–Whitney U | <0.001 | -0.213 | Cliff's delta | <0.05 | TRUE |
| Smoke | categorical | Chi-square | 0.21 | 0.015 | Cramér's V | 0.29 | FALSE |
| Fetal sex | categorical | Chi-square | <0.001 | 0.045 | Cramér's V | <0.05 | TRUE |
| Pre-pregnancy BMI | numeric | Mann–Whitney U | <0.001 | -0.261 | Cliff's delta | <0.001 | TRUE |

| Variable | Type | Test | P value | Effect size | Effect type | Q value | Significant |
|---|---|---|---|---|---|---|---|
| Preexisting diabetes | categorical | Fisher's exact (2x2) | 0.28 | 0.011 | Cramér's V | 0.37 | FALSE |
| HEI - Total Vegetables | numeric | Mann–Whitney U | 0.12 | -0.098 | Cliff's delta | 0.22 | FALSE |
| HEI - Vegetables & Legumes | numeric | Mann–Whitney U | 0.66 | -0.027 | Cliff's delta | 0.72 | FALSE |
| HEI - Total Fruit | numeric | Mann–Whitney U | 0.24 | -0.073 | Cliff's delta | 0.32 | FALSE |
| HEI - Whole Fruit | numeric | Mann–Whitney U | 0.16 | -0.083 | Cliff's delta | 0.25 | FALSE |
| HEI - Whole Grains | numeric | Mann–Whitney U | 0.40 | -0.055 | Cliff's delta | 0.48 | FALSE |
| HEI - Milk | numeric | Mann–Whitney U | 0.45 | 0.049 | Cliff's delta | 0.52 | FALSE |
| HEI - Meat and Beans | numeric | Mann–Whitney U | 0.13 | -0.094 | Cliff's delta | 0.22 | FALSE |
| HEI - Seafood & Plant Protein | numeric | Mann–Whitney U | 0.05 | -0.120 | Cliff's delta | 0.12 | FALSE |
| HEI - Fatty Acid Ratio | numeric | Mann–Whitney U | <0.05 | -0.146 | Cliff's delta | 0.06 | FALSE |
| HEI - Sodium | numeric | Mann–Whitney U | <0.05 | 0.142 | Cliff's delta | 0.07 | FALSE |
| HEI - Refined Grains | numeric | Mann–Whitney U | 0.78 | 0.017 | Cliff's delta | 0.83 | FALSE |
| HEI - SoFAAS Cals | numeric | Mann–Whitney U | <0.05 | -0.172 | Cliff's delta | <0.05 | TRUE |
| HEI - Total Score | numeric | Mann–Whitney U | 0.06 | -0.124 | Cliff's delta | 0.12 | FALSE |
| AHEI - Vegetable servings score | numeric | Mann–Whitney U | 0.47 | -0.047 | Cliff's delta | 0.53 | FALSE |
| AHEI - Fruit servings score | numeric | Mann–Whitney U | 0.18 | -0.088 | Cliff's delta | 0.26 | FALSE |
| AHEI - Whole grain servings score | numeric | Mann–Whitney U | 0.64 | -0.035 | Cliff's delta | 0.71 | FALSE |
| AHEI - Sugary beverages servings score | numeric | Mann–Whitney U | 0.07 | -0.112 | Cliff's delta | 0.13 | FALSE |

| Variable | Type | Test | P value | Effect size | Effect type | Q value | Significant |
|---|---|---|---|---|---|---|---|
| AHEI - Nuts and legumes servings score | numeric | Mann–Whitney U | 0.21 | -0.081 | Cliff's delta | 0.29 | FALSE |
| AHEI - Red meats servings score | numeric | Mann–Whitney U | 0.96 | 0.003 | Cliff's delta | 0.96 | FALSE |
| AHEI - Trans-fat percent score | numeric | Mann–Whitney U | 0.34 | -0.062 | Cliff's delta | 0.43 | FALSE |
| AHEI - DHA & EPA intake score | numeric | Mann–Whitney U | 0.96 | 0.003 | Cliff's delta | 0.96 | FALSE |
| AHEI - Polyunsaturated fat percent score | numeric | Mann–Whitney U | <0.05 | -0.135 | Cliff's delta | 0.08 | FALSE |
| AHEI - Sodium intake score | numeric | Mann–Whitney U | 0.86 | -0.011 | Cliff's delta | 0.88 | FALSE |
| AHEI - Alcoholic drinks score | categorical | Chi-square | <0.001 | 0.056 | Cramér's V | <0.001 | TRUE |
| AHEI - Total Score | numeric | Mann–Whitney U | <0.001 | -0.195 | Cliff's delta | <0.05 | TRUE |
| BMI at visit 1 | numeric | Mann–Whitney U | <0.001 | -0.289 | Cliff's delta | <0.001 | TRUE |
| waist circumference at visit 1 | numeric | Mann–Whitney U | 0.05 | -0.128 | Cliff's delta | 0.11 | FALSE |
| waist over iliac crest at visit 1 | numeric | Mann–Whitney U | <0.001 | -0.297 | Cliff's delta | <0.001 | TRUE |
| Hip circumference at visit 1 | numeric | Mann–Whitney U | <0.001 | -0.314 | Cliff's delta | <0.001 | TRUE |
| Neck circumference at visit 1 | numeric | Mann–Whitney U | <0.001 | -0.315 | Cliff's delta | <0.001 | TRUE |
| SBP at visit 1 | numeric | Mann–Whitney U | <0.001 | -0.316 | Cliff's delta | <0.001 | TRUE |
| DBP at visit 1 | numeric | Mann–Whitney U | <0.05 | -0.177 | Cliff's delta | <0.05 | TRUE |
| GWG at visit 1 | numeric | Mann–Whitney U | 0.15 | -0.093 | Cliff's delta | 0.24 | FALSE |
| GWG at visit 2 | numeric | Mann–Whitney U | <0.05 | -0.209 | Cliff's delta | <0.05 | TRUE |
| SBP at visit 2 | numeric | Mann–Whitney U | 0.14 | -0.096 | Cliff's delta | 0.23 | FALSE |

| Variable | Type | Test | P value | Effect size | Effect type | Q value | Significant |
|---|---|---|---|---|---|---|---|
| DBP at visit 2 | numeric | Mann–Whitney U | 0.18 | -0.088 | Cliff's delta | 0.26 | FALSE |
| GDM | categorical | Fisher's exact (2x2) | 0.39 | 0.010 | Cramér's V | 0.48 | FALSE |
| GA at visit 2 | numeric | Mann–Whitney U | 0.79 | -0.016 | Cliff's delta | 0.83 | FALSE |
| BPD at visit 2 | numeric | Mann–Whitney U | 0.08 | -0.112 | Cliff's delta | 0.16 | FALSE |
| HC at visit 2 | numeric | Mann–Whitney U | <0.05 | -0.154 | Cliff's delta | 0.05 | FALSE |
| AC at visit 2 | numeric | Mann–Whitney U | <0.05 | -0.198 | Cliff's delta | <0.05 | TRUE |
| FL at visit 2 | numeric | Mann–Whitney U | 0.17 | -0.088 | Cliff's delta | 0.26 | FALSE |
| EFW at visit 2 | numeric | Mann–Whitney U | <0.05 | -0.150 | Cliff's delta | 0.05 | FALSE |
| GWG at visit 3 | numeric | Mann–Whitney U | <0.05 | -0.203 | Cliff's delta | <0.05 | TRUE |
| SBP at visit 3 | numeric | Mann–Whitney U | 0.10 | -0.106 | Cliff's delta | 0.19 | FALSE |
| DBP at visit 3 | numeric | Mann–Whitney U | 0.34 | -0.061 | Cliff's delta | 0.43 | FALSE |
| GA at visit 3 | numeric | Mann–Whitney U | 0.23 | -0.079 | Cliff's delta | 0.31 | FALSE |
| BPD at visit 3 | numeric | Mann–Whitney U | <0.001 | -0.431 | Cliff's delta | <0.001 | TRUE |
| HC at visit 3 | numeric | Mann–Whitney U | <0.001 | -0.411 | Cliff's delta | <0.001 | TRUE |
| AC at visit 3 | numeric | Mann–Whitney U | <0.001 | -0.501 | Cliff's delta | <0.001 | TRUE |
| FL at visit 3 | numeric | Mann–Whitney U | <0.001 | -0.330 | Cliff's delta | <0.001 | TRUE |
| EFW at visit 3 | numeric | Mann–Whitney U | <0.05 | -0.169 | Cliff's delta | <0.05 | TRUE |
| AFI - Quadrant 1 at visit 3 | numeric | Mann–Whitney U | <0.001 | -0.305 | Cliff's delta | <0.001 | TRUE |

| Variable | Type | Test | P value | Effect size | Effect type | Q value | Significant |
|---|---|---|---|---|---|---|---|
| AFI - Quadrant 2 at visit 3 | numeric | Mann–Whitney U | <0.05 | -0.202 | Cliff's delta | <0.05 | TRUE |
| AFI - Quadrant 3 at visit 3 | numeric | Mann–Whitney U | <0.001 | -0.473 | Cliff's delta | <0.001 | TRUE |
| AFI - Quadrant 4 at visit 3 | numeric | Mann–Whitney U | <0.001 | -0.550 | Cliff's delta | <0.001 | TRUE |

**Supplementary Table S3. Definition and analytical role of WHO fetal growth chart–based percentile variables.**

| **Variable name** | **Visit** | **nuMoM2b input(s)** | **WHO chart input** | **Derived variable meaning** | **Role in analysis** |
|---|---|---|---|---|---|
| WHO-HC-2 | Visit 2 | Head circumference at Visit 2 + gestational age at scan | HC + GA | WHO fetal growth percentile based on head circumference at Visit 2 | Included in Table 3 benchmark comparison |
| WHO-BPD-2 | Visit 2 | Biparietal diameter at Visit 2 + gestational age at scan | BPD + GA | WHO fetal growth percentile based on biparietal diameter at Visit 2 | Included in Table 3 benchmark comparison |
| WHO-AC-2 | Visit 2 | Abdominal circumference at Visit 2 + gestational age at scan | AC + GA | WHO fetal growth percentile based on abdominal circumference at Visit 2 | Included in Table 3 benchmark comparison |
| WHO-FL-2 | Visit 2 | Femur length at Visit 2 + gestational age at scan | FL + GA | WHO fetal growth percentile based on femur length at Visit 2 | Included in Table 3 benchmark comparison |
| WHO-EFW-2 | Visit 2 | Hadlock-derived estimated fetal weight at Visit 2 + gestational age at scan | EFW + GA | WHO fetal growth percentile based on estimated fetal weight at Visit 2 | Included in Table 3 benchmark comparison; clinically emphasized proxy |
| WHO-HC-3 | Visit 3 | Head circumference at Visit 3 + gestational age at scan | HC + GA | WHO fetal growth percentile based on head circumference at Visit 3 | Included in Table 3 benchmark comparison |
| WHO-BPD-3 | Visit 3 | Biparietal diameter at Visit 3 + gestational age at scan | BPD + GA | WHO fetal growth percentile based on biparietal diameter at Visit 3 | Included in Table 3 benchmark comparison |
| WHO-AC-3 | Visit 3 | Abdominal circumference at Visit 3 + gestational age at scan | AC + GA | WHO fetal growth percentile based on abdominal circumference at Visit 3 | Included in Table 3 benchmark comparison |
| WHO-FL-3 | Visit 3 | Femur length at Visit 3 + | FL + GA | WHO fetal growth percentile based on femur length at Visit 3 | Included in Table 3 benchmark comparison |

| Variable name | Visit | nuMoM2b input(s) | WHO chart input | Derived variable meaning | Role in analysis |
|---|---|---|---|---|---|
| | | gestational age at scan | | | |
| WHO-EFW-3 | Visit 3 | Hadlock-derived estimated fetal weight at Visit 3 + gestational age at scan | EFW + GA | WHO fetal growth percentile based on estimated fetal weight at Visit 3 | Included in Table 3 benchmark comparison; clinically emphasized proxy |

Abbreviations: AC, abdominal circumference; BPD, biparietal diameter; EFW, estimated fetal weight; FL, femur length; GA, gestational age; HC, head circumference; WHO, World Health Organization.

Note: Estimated fetal weight was calculated using the Hadlock formula, and WHO growth-chart percentiles were assigned according to gestational age and the corresponding ultrasound-derived fetal biometric measurement. All 10 WHO-derived percentile variables were evaluated in the benchmark comparison reported in Table 3. These comparisons are descriptive.

### Supplementary Table S4. Discrimination and calibration of the prediction models for LGA sensitivity outcomes.

| Outcome | Model | Event rate | AUROC (95% CI) | AUPRC (95% CI) | Brier score | Scaled Brier score | Calibration intercept | Calibration slope | CITL | O/E |
|---|---|---|---|---|---|---|---|---|---|---|
| LGA90 | Model 1 | 20.7% | 0.620 (0.604–0.637) | 0.285 (0.267–0.306) | 0.1598 | 2.7% | −0.005 | 0.997 | −0.001 | 0.999 |
| LGA90 | Model 2 | 20.7% | 0.654 (0.638–0.670) | 0.324 (0.302–0.348) | 0.1563 | 4.8% | 0.001 | 1.002 | −0.001 | 0.999 |
| LGA90 | Model 3 | 20.7% | 0.731 (0.716–0.745) | 0.414 (0.388–0.441) | 0.1454 | 11.4% | −0.010 | 0.993 | −0.003 | 0.998 |
| LGA90 | Combined model | 20.7% | 0.733 (0.719–0.748) | 0.420 (0.394–0.448) | 0.1448 | 11.8% | −0.008 | 0.995 | −0.003 | 0.998 |
| LGA97 | Model 1 | 7.4% | 0.651 (0.626–0.675) | 0.132 (0.114–0.155) | 0.0670 | 2.0% | −0.022 | 0.992 | −0.002 | 0.998 |
| LGA97 | Model 2 | 7.4% | 0.674 (0.648–0.698) | 0.154 (0.133–0.183) | 0.0662 | 3.2% | 0.014 | 1.007 | −0.002 | 0.998 |
| LGA97 | Model 3 | 7.4% | 0.758 (0.734–0.780) | 0.225 (0.193–0.261) | 0.0631 | 7.7% | −0.034 | 0.987 | −0.006 | 0.995 |

| Outcome | Model | Event rate | AUROC (95% CI) | AUPRC (95% CI) | Brier score | Scaled Brier score | Calibration intercept | Calibration slope | CITL | O/E |
|---|---|---|---|---|---|---|---|---|---|---|
| LGA97 | Combined model | 7.4% | 0.759 (0.736–0.780) | 0.229 (0.195–0.267) | 0.0629 | 7.9% | −0.024 | 0.992 | −0.006 | 0.995 |

Table note: LGA sensitivity outcomes were defined using INTERGROWTH-21st birthweight centiles according to gestational age at delivery and neonatal sex. AUROC and AUPRC were calculated from out-of-fold predictions averaged for each participant across repeated stratified 10-fold cross-validation; 95% confidence intervals were obtained from 1,000 bootstrap resamples of participants. AUPRC was calculated as average precision. Calibration measures were calculated from Platt-recalibrated out-of-fold predictions from one complete run of stratified 10-fold cross-validation, in which each participant had one prediction. The scaled Brier score is one minus the ratio of the Brier score to that of a null model assigning the observed prevalence to all participants (null Brier score 0.1642 for LGA90 and 0.0683 for LGA97); it was calculated from unrounded values. The calibration intercept and slope were estimated by regressing the outcome on the logit of the predicted probability; calibration-in-the-large (CITL) is the intercept with the slope fixed at 1. AUROC: area under the receiver operating characteristic curve; AUPRC: area under the precision–recall curve; CI: confidence interval; LGA: large for gestational age; O/E: observed-to-expected ratio.

### Supplementary Table S5. Discrimination and calibration of the prediction models for the primary macrosomia outcomes.

(a) Discrimination and calibration

| Outcome | Model | Event rate | AUROC (95% CI) | AUPRC (95% CI) | Brier score | Scaled Brier score | Calibration intercept | Calibration slope | CITL | O/E |
|---|---|---|---|---|---|---|---|---|---|---|
| Birthweight >4000 g | Model 1 | 7.9% | 0.628 (0.602–0.651) | 0.120 (0.106–0.139) | 0.0721 | 1.2% | 0.017 | 1.007 | −0.001 | 0.999 |
| Birthweight >4000 g | Model 2 | 7.9% | 0.651 (0.625–0.674) | 0.136 (0.119–0.159) | 0.0715 | 2.1% | 0.039 | 1.017 | −0.001 | 0.999 |
| Birthweight >4000 g | Model 3 | 7.9% | 0.733 (0.709–0.755) | 0.200 (0.176–0.233) | 0.0686 | 6.0% | −0.006 | 0.999 | −0.003 | 0.997 |
| Birthweight >4000 g | Combined model | 7.9% | 0.732 (0.707–0.754) | 0.209 (0.181–0.240) | 0.0686 | 6.0% | −0.002 | 1.001 | −0.003 | 0.997 |
| Birthweight >4500 g | Model 1 | 1.2% | 0.721 (0.670–0.770) | 0.026 (0.017–0.052) | 0.0119 | 0.6% | −0.007 | 0.999 | −0.005 | 0.995 |
| Birthweight >4500 g | Model 2 | 1.2% | 0.727 (0.675–0.766) | 0.037 (0.018–0.068) | 0.0119 | 0.7% | 0.059 | 1.016 | −0.005 | 0.995 |

| Outcome | Model | Event rate | AUROC (95% CI) | AUPRC (95% CI) | Brier score | Scaled Brier score | Calibration intercept | Calibration slope | CITL | O/E |
|---|---|---|---|---|---|---|---|---|---|---|
| Birthweight >4500 g | Model 3 | 1.2% | 0.823 (0.775–0.864) | 0.073 (0.040–0.129) | 0.0116 | 3.3% | −0.046 | 0.992 | −0.018 | 0.983 |
| Birthweight >4500 g | Combined model | 1.2% | 0.821 (0.778–0.866) | 0.066 (0.042–0.107) | 0.0116 | 2.9% | −0.030 | 0.997 | −0.017 | 0.984 |

(b) Paired differences in AUROC between models

| Comparison | Birthweight >4000 g, ΔAUROC (95% CI) | Birthweight >4500 g, ΔAUROC (95% CI) |
|---|---|---|
| Model 2 − Model 1 | 0.023 (0.013 to 0.037) | 0.006 (−0.006 to 0.019) |
| Model 3 − Model 1 | 0.105 (0.081 to 0.128) | 0.102 (0.064 to 0.142) |
| Combined − Model 1 | 0.104 (0.081 to 0.129) | 0.101 (0.061 to 0.139) |
| Model 3 − Model 2 | 0.081 (0.058 to 0.102) | 0.096 (0.058 to 0.134) |
| Combined − Model 2 | 0.081 (0.059 to 0.101) | 0.094 (0.055 to 0.131) |
| Combined − Model 3 | −0.001 (−0.004 to 0.002) | −0.002 (−0.004 to 0.001) |

Table note: AUROC and AUPRC were calculated from out-of-fold predictions averaged for each participant across repeated stratified 10-fold cross-validation; 95% confidence intervals were obtained from 1,000 bootstrap resamples of participants. AUPRC was calculated as average precision. Differences in AUROC were estimated by paired bootstrap resampling, in which the same resampled participants were used for both models. Calibration measures were calculated from Platt-recalibrated out-of-fold predictions from one complete run of stratified 10-fold cross-validation, in which each participant had one prediction. The scaled Brier score is one minus the ratio of the Brier score to that of a null model assigning the observed prevalence to all participants (null Brier score 0.0730 for birthweight >4000 g and 0.0119 for birthweight >4500 g); it was calculated from unrounded values. The calibration intercept and slope were estimated by regressing the outcome on the logit of the predicted probability; calibration-in-the-large (CITL) is the intercept with the slope fixed at 1. AUROC: area under the receiver operating characteristic curve; AUPRC: area under the precision–recall curve; CI: confidence interval; O/E: observed-to-expected ratio.

**Supplementary Table S6. Comparison of included pregnancies and sequentially excluded pregnancies.**

| Characteristic | Included analytic cohort N=6,371 | Excluded: missing diet N=1,349 | P value | Excluded: incomplete Visit 2/3 ultrasound N=814 | P value |
|---|---|---|---|---|---|
| Maternal age, years | 27.44 (5.55) | 25.20 (5.76) | <0.001 | 25.63 (5.40) | <0.001 |
| Non-Hispanic White | 4,081 (64.1) | 645 (47.8) | <0.001 | 496 (60.9) | 0.081 |
| Non-Hispanic Black | 694 (10.9) | 332 (24.6) | <0.001 | 87 (10.7) | 0.859 |
| Hispanic | 1,022 (16.0) | 242 (17.9) | 0.084 | 171 (21.0) | <0.001 |
| Asian/Other race or ethnicity | 574 (9.0) | 130 (9.6) | 0.479 | 60 (7.4) | 0.117 |
| Income | 454.99 (291.98) | 359.06 (305.60) | <0.001 | 346.81 (274.31) | <0.001 |
| Smoking before pregnancy | 1,078 (16.9) | 317 (23.5) | <0.001 | 132 (16.2) | 0.613 |
| Education level: ≤HS grad | 1,097 (17.2) | 397 (29.4) | <0.001 | 186 (22.9) | <0.001 |
| Education level: Some college or Assoc/Tech degree | 1,778 (27.9) | 470 (34.8) | <0.001 | 288 (35.4) | <0.001 |
| Education level: Completed college | 1,887 (29.6) | 255 (18.9) | <0.001 | 223 (27.4) | 0.190 |
| Education level: Degree work beyond college | 1,609 (25.3) | 227 (16.8) | <0.001 | 117 (14.4) | <0.001 |
| Family history of diabetes | 1,369 (21.5) | 286 (21.2) | 0.815 | 164 (20.1) | 0.379 |
| Preexisting diabetes | 88 (1.4) | 28 (2.1) | 0.057 | 15 (1.8) | 0.297 |
| Gestational diabetes | 284 (4.5) | 60 (4.4) | 0.987 | 22 (2.7) | 0.020 |
| Male fetal sex | 3,244 (50.9) | 707 (52.4) | 0.320 | 433 (53.2) | 0.221 |
| Visit-1 BMI, kg/m$^2$ | 26.30 (6.25) | 26.96 (6.66) | 0.001 | 26.11 (6.34) | 0.440 |
| Early gestational weight gain | 2.12 (3.78) | 2.08 (4.71) | 0.776 | 2.42 (3.84) | 0.040 |
| Visit-1 waist circumference | 82.66 (13.10) | 84.80 (14.24) | <0.001 | 83.75 (13.31) | 0.029 |
| Visit-1 waist over iliac crest | 94.83 (14.21) | 95.82 (15.22) | 0.034 | 94.80 (14.04) | 0.958 |
| Visit-1 hip circumference | 104.13 (12.71) | 105.03 (14.15) | 0.038 | 104.36 (13.08) | 0.649 |
| Visit-1 neck circumference | 32.81 (3.01) | 33.20 (3.09) | <0.001 | 33.12 (2.90) | 0.009 |
| Visit-1 systolic blood pressure | 109.05 (10.73) | 109.97 (11.42) | 0.008 | 109.28 (11.62) | 0.597 |
| Visit-1 diastolic blood pressure | 67.08 (8.29) | 66.36 (8.58) | 0.006 | 67.43 (8.60) | 0.278 |
| Gestational age at delivery, weeks | 38.51 (1.83) | 38.19 (2.38) | <0.001 | 37.86 (3.26) | <0.001 |
| Preterm birth <37 weeks | 880 (13.8) | 249 (18.5) | <0.001 | 151 (18.6) | <0.001 |

| Characteristic | Included analytic cohort N=6,371 | Excluded: missing diet N=1,349 | P value | Excluded: incomplete Visit 2/3 ultrasound N=814 | P value |
|---|---|---|---|---|---|
| Birthweight, g | 3,296.44 (534.50) | 3,194.24 (586.57) | <0.001 | 3,165.25 (711.09) | <0.001 |
| Birthweight >4000 g | 505 (7.9) | 82 (6.1) | 0.029 | 44 (5.4) | 0.015 |
| Birthweight >4500 g | 77 (1.2) | 6 (0.4) | 0.015 | 4 (0.5) | 0.072 |

**Supplementary Table S7. Threshold-based operating characteristics of the recalibrated models at the upper quintile and upper decile of predicted risk.**

| Outcome | Model | Risk stratum | Predicted-risk cutoff | TP | FN | FP | TN | Sensitivity (%) | Specificity (%) | PPV (%) | NPV (%) |
|---|---|---|---|---|---|---|---|---|---|---|---|
| Birthweight >4000 g | Model 1 | Upper quintile (top 20%) | 10.0% | 166 | 339 | 1109 | 4757 | 32.9 | 81.1 | 13.0 | 93.3 |
| Birthweight >4000 g | Model 1 | Upper decile (top 10%) | 11.8% | 90 | 415 | 548 | 5318 | 17.8 | 90.7 | 14.1 | 92.8 |
| Birthweight >4000 g | Model 2 | Upper quintile (top 20%) | 10.5% | 180 | 325 | 1095 | 4771 | 35.6 | 81.3 | 14.1 | 93.6 |
| Birthweight >4000 g | Model 2 | Upper decile (top 10%) | 12.8% | 102 | 403 | 536 | 5330 | 20.2 | 90.9 | 16.0 | 93.0 |
| Birthweight >4000 g | Model 3 | Upper quintile (top 20%) | 12.2% | 243 | 262 | 1032 | 4834 | 48.1 | 82.4 | 19.1 | 94.9 |
| Birthweight >4000 g | Model 3 | Upper decile (top 10%) | 16.1% | 148 | 357 | 490 | 5376 | 29.3 | 91.6 | 23.2 | 93.8 |
| Birthweight >4000 g | Combined model | Upper quintile (top 20%) | 12.2% | 239 | 266 | 1036 | 4830 | 47.3 | 82.3 | 18.7 | 94.8 |
| Birthweight >4000 g | Combined model | Upper decile (top 10%) | 16.2% | 147 | 358 | 491 | 5375 | 29.1 | 91.6 | 23.0 | 93.8 |

| Outcome | Model | Risk stratum | Predicted-risk cutoff | TP | FN | FP | TN | Sensitivity (%) | Specificity (%) | PPV (%) | NPV (%) |
|---|---|---|---|---|---|---|---|---|---|---|---|
| Birthweight >4500 g | Model 1 | Upper quintile (top 20%) | 1.7% | 35 | 42 | 1240 | 5054 | 45.5 | 80.3 | 2.7 | 99.2 |
| Birthweight >4500 g | Model 1 | Upper decile (top 10%) | 2.4% | 23 | 54 | 615 | 5679 | 29.9 | 90.2 | 3.6 | 99.1 |
| Birthweight >4500 g | Model 2 | Upper quintile (top 20%) | 1.8% | 37 | 40 | 1238 | 5056 | 48.1 | 80.3 | 2.9 | 99.2 |
| Birthweight >4500 g | Model 2 | Upper decile (top 10%) | 2.4% | 25 | 52 | 613 | 5681 | 32.5 | 90.3 | 3.9 | 99.1 |
| Birthweight >4500 g | Model 3 | Upper quintile (top 20%) | 1.7% | 56 | 21 | 1219 | 5075 | 72.7 | 80.6 | 4.4 | 99.6 |
| Birthweight >4500 g | Model 3 | Upper decile (top 10%) | 3.0% | 36 | 41 | 602 | 5692 | 46.8 | 90.4 | 5.6 | 99.3 |
| Birthweight >4500 g | Combined model | Upper quintile (top 20%) | 1.8% | 54 | 23 | 1221 | 5073 | 70.1 | 80.6 | 4.2 | 99.5 |
| Birthweight >4500 g | Combined model | Upper decile (top 10%) | 3.0% | 35 | 42 | 603 | 5691 | 45.5 | 90.4 | 5.5 | 99.3 |
| LGA90 | Model 1 | Upper quintile (top 20%) | 24.8% | 398 | 921 | 877 | 4175 | 30.2 | 82.6 | 31.2 | 81.9 |
| LGA90 | Model 1 | Upper decile (top 10%) | 29.1% | 219 | 1100 | 419 | 4633 | 16.6 | 91.7 | 34.3 | 80.8 |
| LGA90 | Model 2 | Upper quintile (top 20%) | 26.5% | 443 | 876 | 832 | 4220 | 33.6 | 83.5 | 34.7 | 82.8 |

| Outcome | Model | Risk stratum | Predicted-risk cutoff | TP | FN | FP | TN | Sensitivity (%) | Specificity (%) | PPV (%) | NPV (%) |
|---|---|---|---|---|---|---|---|---|---|---|---|
| LGA90 | Model 2 | Upper decile (top 10%) | 32.1% | 255 | 1064 | 383 | 4669 | 19.3 | 92.4 | 40.0 | 81.4 |
| LGA90 | Model 3 | Upper quintile (top 20%) | 31.3% | 554 | 765 | 721 | 4331 | 42.0 | 85.7 | 43.5 | 85.0 |
| LGA90 | Model 3 | Upper decile (top 10%) | 38.6% | 313 | 1006 | 325 | 4727 | 23.7 | 93.6 | 49.1 | 82.5 |
| LGA90 | Combined model | Upper quintile (top 20%) | 31.5% | 560 | 759 | 715 | 4337 | 42.5 | 85.8 | 43.9 | 85.1 |
| LGA90 | Combined model | Upper decile (top 10%) | 38.7% | 322 | 997 | 316 | 4736 | 24.4 | 93.7 | 50.5 | 82.6 |
| LGA97 | Model 1 | Upper quintile (top 20%) | 9.1% | 178 | 292 | 1097 | 4804 | 37.9 | 81.4 | 14.0 | 94.3 |
| LGA97 | Model 1 | Upper decile (top 10%) | 11.5% | 111 | 359 | 527 | 5374 | 23.6 | 91.1 | 17.4 | 93.7 |
| LGA97 | Model 2 | Upper quintile (top 20%) | 9.4% | 194 | 276 | 1081 | 4820 | 41.3 | 81.7 | 15.2 | 94.6 |
| LGA97 | Model 2 | Upper decile (top 10%) | 12.2% | 120 | 350 | 518 | 5383 | 25.5 | 91.2 | 18.8 | 93.9 |
| LGA97 | Model 3 | Upper quintile (top 20%) | 11.4% | 249 | 221 | 1026 | 4875 | 53.0 | 82.6 | 19.5 | 95.7 |
| LGA97 | Model 3 | Upper decile (top 10%) | 15.7% | 164 | 306 | 474 | 5427 | 34.9 | 92.0 | 25.7 | 94.7 |
| LGA97 | Combined | Upper quintile | 11.2% | 245 | 225 | 1030 | 4871 | 52.1 | 82.5 | 19.2 | 95.6 |

| Outcome | Model | Risk stratum | Predicted-risk cutoff | TP | FN | FP | TN | Sensitivity (%) | Specificity (%) | PPV (%) | NPV (%) |
|---|---|---|---|---|---|---|---|---|---|---|---|
| | model | (top 20%) | | | | | | | | | |
| LGA97 | Combined model | Upper decile (top 10%) | 15.6% | 167 | 303 | 471 | 5430 | 35.5 | 92.0 | 26.2 | 94.7 |

Table note: Threshold-based operating characteristics were calculated from Platt-recalibrated out-of-fold predicted probabilities from one complete run of stratified 10-fold cross-validation, in which each participant had one prediction. Predictions from all folds were ranked together, and the 1,275 pregnancies (upper quintile) and 638 pregnancies (upper decile) with the highest predicted risk were classified as screen-positive. The predicted-risk cutoff is the lowest recalibrated predicted probability among screen-positive pregnancies. These fixed risk strata describe screening yield and do not represent recommended clinical intervention thresholds. TP: true positive; FN: false negative; FP: false positive; TN: true negative; PPV: positive predictive value; NPV: negative predictive value.

### Supplementary Table S8. Sensitivity comparison of logistic regression and non-linear classifiers for primary macrosomia outcomes.

| Outcome | Model | Classifier | AUROC (95% CI) | AUPRC (95% CI) |
|---|---|---|---|---|
| Birthweight >4000 g | Model 1 | Logistic regression | 0.628 (0.602–0.651) | 0.120 (0.106–0.139) |
| Birthweight >4000 g | Model 1 | Random forest | 0.634 (0.608–0.660) | 0.118 (0.105–0.136) |
| Birthweight >4000 g | Model 1 | XGBoost | 0.626 (0.602–0.652) | 0.118 (0.105–0.136) |
| Birthweight >4000 g | Model 1 | Gradient boosting | 0.615 (0.590–0.640) | 0.115 (0.102–0.132) |
| Birthweight >4000 g | Model 2 | Logistic regression | 0.651 (0.625–0.674) | 0.136 (0.119–0.159) |
| Birthweight >4000 g | Model 2 | Random forest | 0.650 (0.626–0.676) | 0.131 (0.114–0.153) |
| Birthweight >4000 g | Model 2 | XGBoost | 0.634 (0.609–0.661) | 0.131 (0.115–0.155) |
| Birthweight >4000 g | Model 2 | Gradient boosting | 0.640 (0.616–0.667) | 0.133 (0.116–0.155) |
| Birthweight >4000 g | Model 3 | Logistic regression | 0.733 (0.709–0.755) | 0.200 (0.176–0.233) |
| Birthweight >4000 g | Model 3 | Random forest | 0.731 (0.709–0.755) | 0.207 (0.181–0.242) |
| Birthweight >4000 g | Model 3 | XGBoost | 0.734 (0.712–0.758) | 0.201 (0.177–0.235) |
| Birthweight >4000 g | Model 3 | Gradient boosting | 0.736 (0.714–0.760) | 0.208 (0.182–0.243) |
| Birthweight >4000 g | Combined model | Logistic regression | 0.732 (0.707–0.754) | 0.209 (0.181–0.240) |

| Outcome | Model | Classifier | AUROC (95% CI) | AUPRC (95% CI) |
|---|---|---|---|---|
| Birthweight >4000 g | Combined model | Random forest | 0.735 (0.712–0.759) | 0.208 (0.181–0.243) |
| Birthweight >4000 g | Combined model | XGBoost | 0.730 (0.708–0.754) | 0.196 (0.172–0.227) |
| Birthweight >4000 g | Combined model | Gradient boosting | 0.738 (0.717–0.761) | 0.201 (0.177–0.233) |
| Birthweight >4500 g | Model 1 | Logistic regression | 0.721 (0.670–0.770) | 0.026 (0.017–0.052) |
| Birthweight >4500 g | Model 1 | Random forest | 0.693 (0.632–0.754) | 0.023 (0.017–0.032) |
| Birthweight >4500 g | Model 1 | XGBoost | 0.667 (0.599–0.733) | 0.020 (0.014–0.027) |
| Birthweight >4500 g | Model 1 | Gradient boosting | 0.681 (0.618–0.738) | 0.021 (0.016–0.029) |
| Birthweight >4500 g | Model 2 | Logistic regression | 0.727 (0.675–0.766) | 0.037 (0.018–0.068) |
| Birthweight >4500 g | Model 2 | Random forest | 0.704 (0.638–0.769) | 0.025 (0.020–0.031) |
| Birthweight >4500 g | Model 2 | XGBoost | 0.673 (0.609–0.733) | 0.023 (0.018–0.031) |
| Birthweight >4500 g | Model 2 | Gradient boosting | 0.711 (0.647–0.768) | 0.025 (0.020–0.033) |
| Birthweight >4500 g | Model 3 | Logistic regression | 0.823 (0.775–0.864) | 0.073 (0.040–0.129) |
| Birthweight >4500 g | Model 3 | Random forest | 0.804 (0.757–0.852) | 0.066 (0.042–0.116) |
| Birthweight >4500 g | Model 3 | XGBoost | 0.782 (0.728–0.836) | 0.066 (0.041–0.117) |
| Birthweight >4500 g | Model 3 | Gradient boosting | 0.798 (0.743–0.849) | 0.065 (0.042–0.108) |
| Birthweight >4500 g | Combined model | Logistic regression | 0.821 (0.778–0.866) | 0.066 (0.042–0.107) |
| Birthweight >4500 g | Combined model | Random forest | 0.820 (0.774–0.866) | 0.058 (0.038–0.096) |
| Birthweight >4500 g | Combined model | XGBoost | 0.778 (0.724–0.831) | 0.061 (0.039–0.108) |
| Birthweight >4500 g | Combined model | Gradient boosting | 0.788 (0.735–0.838) | 0.063 (0.039–0.114) |

Table note. Logistic-regression rows correspond to the primary models (Table S5). The non-linear classifiers used the same predictor sets as the logistic regression models (42, 51, 56, and 65 predictors) and were evaluated in one run of stratified 10-fold cross-validation, so that each participant had one out-of-fold prediction. For all classifiers, hyperparameters were selected within each outer training fold by randomized search with inner 3-fold cross-validation optimizing average precision; the validation fold was not used for tuning. Class imbalance was handled using fold-specific sample weights, with the negative class assigned a weight of 1 and the positive class weighted by the square root of the negative-to-positive sample ratio. The search spaces were as follows. Logistic regression (20 randomized-search draws; elastic-net penalty, SAGA solver, maximum 50,000 iterations, tolerance $1\times10^{-4}$): C, 0.01, 0.05, 0.1, 0.3, and 1; L1 ratio, 0.1, 0.3, 0.5, 0.7, and 0.9. Random forest (12 randomized-search draws; bootstrap sampling): number of trees, 300, 500, and 800; maximum depth, none, 3, 5, 8, and 12; minimum samples per leaf, 5, 10, 20, and 40; minimum samples per split, 2, 10, 20, and 50; maximum features, square root, 0.30, 0.50, and 0.80. XGBoost (18 draws): number of trees, 200, 400, 600, and 900; learning rate, 0.01, 0.03, 0.05, 0.08, and 0.10; maximum depth, 1–4; minimum child weight, 1, 5, 10, and 20; subsample, 0.60, 0.75, 0.90, and 1.00; column subsample per tree, 0.60, 0.75, 0.90, and 1.00; gamma, 0, 0.1, 0.5, 1.0,

5, and 10; L1 regularization, 0, 0.1, 0.5, and 1.0; L2 regularization, 1, 3, 5, and 10. Gradient boosting (12 draws): number of trees, 100, 200, 300, and 500; learning rate, 0.01, 0.03, 0.05, 0.08, and 0.10; maximum depth, 1–3; minimum samples per leaf, 5, 10, 20, and 40; subsample, 0.60, 0.80, and 1.00; maximum features, none, square root, 0.50, and 0.80. For the non-linear classifiers, 95% confidence intervals were obtained from 2,000 bootstrap resamples of participants; the logistic-regression estimates and confidence intervals were taken from Table S5. AUPRC was calculated as average precision. AUROC: area under the receiver operating characteristic curve; AUPRC: area under the precision–recall curve.

### Supplementary Table S9. Missingness of the predictors included in the prediction models before k-nearest-neighbor imputation.

| Predictor | Visit | Missing, % |
|---|---:|---:|
| Age | Visit 1 | 0.4 |
| Race and ethnicity (four indicators) | Visit 1 | 0.0 |
| Income level | Visit 1 | 16.3 |
| Smoking in the 3 months before pregnancy | Visit 1 | 0.0 |
| Pre-pregnancy BMI | Visit 1 | 1.3 |
| Preexisting diabetes mellitus | Visit 1 | 0.0 |
| Healthy Eating Index-2010 and Alternative Healthy Eating Index-2010 component and total scores (25 predictors) | Visit 1 | 0.0 |
| BMI at Visit 1 | Visit 1 | 2.0 |
| Gestational weight gain at Visit 1 | Visit 1 | 1.6 |
| Waist circumference at Visit 1 | Visit 1 | 2.0 |
| Waist over iliac crest at Visit 1 | Visit 1 | 2.0 |
| Hip circumference at Visit 1 | Visit 1 | 2.1 |
| Neck circumference at Visit 1 | Visit 1 | 12.1 |
| Systolic blood pressure at Visit 1 | Visit 1 | 1.4 |
| Diastolic blood pressure at Visit 1 | Visit 1 | 1.4 |
| Gestational weight gain at Visit 2 | Visit 2 | 2.2 |
| Systolic blood pressure at Visit 2 | Visit 2 | 1.4 |
| Diastolic blood pressure at Visit 2 | Visit 2 | 1.4 |
| Gestational age at ultrasound, Visit 2 | Visit 2 | 0.5 |
| Biparietal diameter, Visit 2 | Visit 2 | 0.0 |
| Head circumference, Visit 2 | Visit 2 | 0.0 |
| Abdominal circumference, Visit 2 | Visit 2 | 0.2 |
| Femur length, Visit 2 | Visit 2 | 0.0 |
| Estimated fetal weight, Visit 2 | Visit 2 | 0.2 |
| Gestational weight gain at Visit 3 | Visit 3 | 2.1 |
| Systolic blood pressure at Visit 3 | Visit 3 | 1.3 |
| Diastolic blood pressure at Visit 3 | Visit 3 | 1.3 |
| Gestational age at ultrasound, Visit 3 | Visit 3 | 0.4 |
| Biparietal diameter, Visit 3 | Visit 3 | 0.0 |
| Head circumference, Visit 3 | Visit 3 | 0.0 |
| Abdominal circumference, Visit 3 | Visit 3 | 0.0 |
| Femur length, Visit 3 | Visit 3 | 0.0 |
| Amniotic fluid index, quadrant 1 | Visit 3 | 7.4 |
| Amniotic fluid index, quadrant 2 | Visit 3 | 7.2 |
| Amniotic fluid index, quadrant 3 | Visit 3 | 7.1 |
| Amniotic fluid index, quadrant 4 | Visit 3 | 8.0 |
| Estimated fetal weight, Visit 3 | Visit 3 | 0.0 |
| Estimated fetal weight percentile reported in nuMoM2b, Visit 3 | Visit 3 | 0.5 |

Note: Missingness was calculated in the final analytic cohort (N=6,371) before k-nearest-neighbor imputation. The table includes all predictors used in the prediction models, as defined in Table 2 of the main manuscript. Residual missingness was handled by k-nearest-neighbor imputation fitted within the training data of each cross-validation split. Variables that were not used as predictors (fetal sex, gestational diabetes mellitus, the Visit-2 reported estimated fetal weight percentile, and uterine artery Doppler measurements) are not shown. Percentages are rounded to one decimal place. BMI: body mass index.

### Supplementary Table S10. Comparison of WHO and INTERGROWTH-21st fetal ultrasound percentile comparators for primary macrosomia outcomes.

| Outcome | Visit | Measure | WHO AUROC | INTERGROWTH-21st AUROC (95% CI) |
|---|---|---|---|---|
| Birthweight >4000 g | Visit 2 | BPD | 0.570 | 0.572 (0.547–0.599) |
| Birthweight >4000 g | Visit 2 | HC | 0.580 | 0.581 (0.558–0.605) |
| Birthweight >4000 g | Visit 2 | AC | 0.590 | 0.589 (0.564–0.614) |
| Birthweight >4000 g | Visit 2 | FL | 0.540 | 0.548 (0.523–0.574) |
| Birthweight >4000 g | Visit 2 | EFW | 0.570 | 0.609 (0.577–0.640)[a] |
| Birthweight >4000 g | Visit 3 | BPD | 0.650 | 0.660 (0.634–0.685) |
| Birthweight >4000 g | Visit 3 | HC | 0.650 | 0.664 (0.638–0.688) |
| Birthweight >4000 g | Visit 3 | AC | 0.700 | 0.707 (0.685–0.730) |
| Birthweight >4000 g | Visit 3 | FL | 0.610 | 0.610 (0.584–0.636) |
| Birthweight >4000 g | Visit 3 | EFW | 0.680 | 0.694 (0.669–0.717) |
| Birthweight >4500 g | Visit 2 | BPD | 0.580 | 0.578 (0.513–0.646) |
| Birthweight >4500 g | Visit 2 | HC | 0.620 | 0.619 (0.548–0.683) |
| Birthweight >4500 g | Visit 2 | AC | 0.640 | 0.639 (0.581–0.696) |
| Birthweight >4500 g | Visit 2 | FL | 0.540 | 0.551 (0.481–0.618) |
| Birthweight >4500 g | Visit 2 | EFW | 0.600 | 0.628 (0.545–0.713)[a] |
| Birthweight >4500 g | Visit 3 | BPD | 0.740 | 0.752 (0.694–0.805) |
| Birthweight >4500 g | Visit 3 | HC | 0.740 | 0.747 (0.685–0.806) |
| Birthweight >4500 g | Visit 3 | AC | 0.790 | 0.799 (0.745–0.846) |
| Birthweight >4500 g | Visit 3 | FL | 0.680 | 0.690 (0.628–0.749) |
| Birthweight >4500 g | Visit 3 | EFW | 0.770 | 0.787 (0.734–0.834) |

Note. Fetal ultrasound percentiles were evaluated as single-predictor chart-based comparators for subsequent macrosomia. INTERGROWTH-21st values are shown as AUROC with 95% confidence intervals from 2,000 bootstrap resamples. INTERGROWTH-21st fetal growth percentiles were derived according to the INTERGROWTH-21st fetal growth standards [28], and estimated fetal weight percentiles according to the INTERGROWTH-21st standards for Hadlock-based estimated fetal weight [29]. [a] The INTERGROWTH-21st standard for Hadlock-based estimated fetal weight applies from 18 weeks of gestation (126–287 days). Because 39.8% of the Visit-2 scans were performed before 18 weeks, this percentile could be derived for 3,834 pregnancies (320 with birthweight >4000 g and 48 with birthweight >4500 g). AUROC: area under the receiver operating characteristic curve; WHO: World Health Organization; BPD: biparietal diameter; HC: head circumference; AC: abdominal circumference; FL: femur length; EFW: estimated fetal weight.

### Supplementary Table S11. TRIPOD+AI 2024 checklist.

| Item | Checklist item | Reported in manuscript | Location | Comments |
|---|---|---|---|---|
| 1 | Title identifies the study as developing and internally validating a prediction model, the target population, and outcome. | Yes | Title page | The title states development and internal validation of prediction models for macrosomia in nulliparous singleton pregnancies. |
| 2 | Abstract summarizes objectives, data source, participants, predictors, outcomes, analytical methods, results, and conclusions. | Yes | Abstract | The abstract reports the nuMoM2b cohort, sample size, predictors, outcomes, logistic regression models, internal validation, comparison with fetal growth chart–based benchmarks, and main performance results. |
| 3a | Background explains the clinical context and rationale for prediction modeling. | Yes | Introduction | The introduction describes macrosomia-related clinical risks and the need for earlier risk stratification. |
| 3b | Target population, intended purpose, and clinical context of model use are described. | Yes | Introduction; Discussion, Sections 4.1 and 4.3 | The intended context is antenatal risk stratification for macrosomia in nulliparous singleton pregnancies. |
| 3c | Relevant population groups or sociodemographic considerations are described where applicable. | Partly | Section 2.3; Table 1; Supplementary Table S6; Discussion, Section 4.4 | Sociodemographic predictors such as race/ethnicity and income were included and summarized. Potential selection during cohort derivation was evaluated in Supplementary Table S6. Formal fairness or subgroup performance analyses were not performed. |
| 4 | Study objectives specify model development and validation purpose. | Yes | End of Introduction | The study objective states development and internal validation of visit-updated prediction models for birthweight >4000 g and >4500 g. |

| Item | Checklist item | Reported in manuscript | Location | Comments |
|---|---|---|---|---|
| 5a | Data source and rationale are described. | Yes | Section 2.1 | The study is a secondary analysis of the prospective nuMoM2b cohort. |
| 5b | Key dates of data collection are reported. | Yes | Section 2.1 | Recruitment period and scheduled visit windows are described. |
| 6a | Study setting is described. | Yes | Section 2.1 | Participants were recruited from hospitals affiliated with eight clinical centers in the United States. |
| 6b | Eligibility criteria and participant selection are described. | Yes | Section 2.1; Section 3.1; Supplementary Figure S4 | Inclusion and exclusion criteria are described, including availability of periconceptional diet, Visit 2 and Visit 3 ultrasound data, birth outcome data, and live-birth status. Participant flow is shown in Supplementary Figure S4. |
| 6c | Treatments or interventions are described if relevant. | Not applicable | — | This was an observational secondary analysis; no treatment assignment or intervention was part of the prediction model study. |
| 7 | Data preparation, preprocessing, and quality checking are described. | Yes | Sections 2.3–2.5; Supplementary Methods; Supplementary Tables S3 and S9 | Predictor construction, ultrasound-derived percentile variables, missing-data handling, imputation, scaling, and cross-validation preprocessing are described. WHO fetal growth chart–based percentile variables are defined in Supplementary Table S3, and predictor-level missingness is reported in Supplementary Table S9. |
| 8a | Outcome definition, time horizon, assessment, and rationale are described. | Yes | Section 2.2; Section 3.1; Supplementary Table S4 | Primary outcomes were birthweight >4000 g and >4500 g. LGA90 and LGA97 were used as sensitivity outcomes and are reported in Supplementary Table S4. |
| 8b | Qualifications of outcome assessors are described if subjective assessment is required. | Not applicable | Section 2.2 | Outcomes were based on recorded birthweight, gestational age at delivery, and neonatal sex, not subjective adjudication. |
| 8c | Blinding of outcome assessment is described if relevant. | Not applicable | — | Outcome assessment was based on recorded clinical birth data; no subjective blinded assessment was required. |

| Item | Checklist item | Reported in manuscript | Location | Comments |
|---|---|---|---|---|
| 9a | Initial predictor selection and rationale are described. | Yes | Section 2.3 | Candidate predictors were prespecified according to clinical availability during antenatal care and prior evidence linking them to fetal overgrowth. |
| 9b | Predictors are clearly defined, including how and when they were measured. | Yes | Section 2.3; Table 2; Supplementary Methods; Supplementary Table S3 | All predictors in each model are listed in Table 2 together with their time of availability. Maternal characteristics, anthropometry, diet indices, and ultrasound predictors are described in Section 2.3. Fetal sex and gestational diabetes mellitus were not used as predictors. WHO fetal growth chart–based percentile variables are further defined in Supplementary Table S3. |
| 9c | Qualifications of predictor assessors are described if subjective interpretation is required. | Partly | Section 2.1; Section 2.3 | Most predictors were routinely collected clinical, anthropometric, ultrasound, or questionnaire variables. Ultrasound measurements followed the standardized nuMoM2b visit structure. |
| 10 | Sample size is explained and justified in the prediction-model context. | Partly | Section 2.6; Section 3.1; Supplementary Table S13 | The sample size was determined by the available nuMoM2b data. The number of candidate predictors and events per candidate predictor are reported for each model and outcome. Because of the limited number of events for birthweight >4500 g, model complexity was additionally assessed (Supplementary Table S13). No a priori sample-size calculation was performed. |
| 11 | Missing data handling is described, including reasons for omitting data. | Yes | Sections 2.1 and 2.5; Supplementary Figure S4; Supplementary Tables S6 and S9 | Cohort-level exclusions are shown in Supplementary Figure S4. Included versus excluded pregnancies are compared in Supplementary Table S6. Predictor-level missingness before imputation is reported in Supplementary Table S9, and k-nearest-neighbor imputation is described in Section 2.5. |

| Item | Checklist item | Reported in manuscript | Location | Comments |
|---|---|---|---|---|
| 12a | Data use for model development and internal validation is described. | Yes | Section 2.5 | Repeated stratified 10-fold cross-validation with 1000 repetitions was used for internal validation. Out-of-fold predictions were averaged at the participant level, and confidence intervals were obtained by bootstrap resampling of participants. |
| 12b | Predictor handling, transformation, scaling, or standardization is described. | Yes | Section 2.5 | Imputation and scaling were performed within the training data only to avoid information leakage. |
| 12c | Model type, rationale, model-building steps, hyperparameter tuning, and internal validation are described. | Yes | Section 2.5; Supplementary Table S8 | Logistic regression was selected as the primary modeling approach. Regularization, class weighting, hyperparameter tuning, Platt recalibration, and internal validation are described in Section 2.5; hyperparameter search spaces are provided in the note to Supplementary Table S8. Non-linear classifier sensitivity comparisons are reported in Supplementary Table S8. |
| 12d | Handling of heterogeneity across clusters or centers is described if performed. | Not performed | — | Formal center-or cluster-specific model-performance heterogeneity was not evaluated. |
| 12e | Performance measures and plots are specified. | Yes | Section 2.5; Section 3.2; Figure 1; Supplementary Figures S1–S3 and S5; Supplementary Tables S4, S5, S7, S8, and S13 | AUROC, AUPRC, paired differences in AUROC, Brier score, scaled Brier score, calibration intercept, calibration slope, calibration-in-the-large, O/E ratio, reliability plots, LGA sensitivity performance, threshold-based operating characteristics, decision curves, and non-linear model sensitivity comparisons are reported. |
| 12f | Model updating is described if performed. | Not applicable | — | No external model updating, recalibration, or refitting after external validation was performed. In the limited evaluation in the MMC cohort, the nuMoM2b coefficients and intercept |

| Item | Checklist item | Reported in manuscript | Location | Comments |
|---|---|---|---|---|
| | | | | were applied without updating. |
| 12g | Calculation of model predictions for evaluation is described. | Yes | Section 2.5; Supplementary Tables S4, S5, S7, and S8 | Out-of-fold predicted probabilities were averaged at the participant level for discrimination analyses. For the main model evaluations, calibration, threshold-based, and decision-curve analyses used Platt-recalibrated out-of-fold predictions from one complete run of 10-fold cross-validation, with one prediction per participant. |
| 13 | Class imbalance handling and recalibration are described. | Yes | Section 2.5; Supplementary Tables S4–S5; Supplementary Figures S2–S3 | Class-weighted logistic regression was used, followed by Platt recalibration fitted within the training folds. Calibration summaries are reported for primary outcomes and LGA sensitivity outcomes. |
| 14 | Model fairness approaches are described if performed. | Not performed | Section 2.3; Table 1; Supplementary Table S6; Discussion, Section 4.4 | Race/ethnicity and income were included as predictors and summarized. Selection during cohort derivation was assessed in Supplementary Table S6. Formal fairness or subgroup performance analyses were not performed. |
| 15 | Model output and thresholds are described. | Yes | Section 2.5; Section 3.2; Supplementary Table S7 | Model output was recalibrated predicted probability. Upper-quintile and upper-decile risk strata were evaluated as fixed risk strata in Supplementary Table S7; they do not represent recommended intervention thresholds. |
| 16 | Differences between development and evaluation data are identified. | Yes | Sections 2.6, 3.2, and 4.4; Supplementary Table S12 | Performance of the full models was assessed by internal validation only. A reduced early-pregnancy model with six harmonizable predictors was evaluated in an independent Dutch cohort (MMC). Differences between the cohorts are described: the MMC data included multiparous women and repeated deliveries, and only six predictors could be harmonized. |

| Item | Checklist item | Reported in manuscript | Location | Comments |
|---|---|---|---|---|
| 17 | Ethical approval and informed consent are reported. | Yes | Ethics Statement | Original nuMoM2b informed consent and institutional approvals for this secondary analysis are reported. The ethics committee decision (waiver of formal ethical approval) for the retrospective use of the MMC data is also reported. |
| 18a | Funding source and funder role are reported. | Yes | Funding Information | Funding source and funder role are stated. |
| 18b | Conflicts of interest are reported. | Yes | Conflict of Interest Statement | Authors declare no conflicts of interest. |
| 18c | Protocol availability is stated. | Partly | Ethics Statement; Data Availability Statement | The secondary analysis was approved by relevant ethics boards. A separate public analysis protocol was not reported. |
| 18d | Study registration is reported if applicable. | Not applicable | — | This secondary analysis was not registered as a clinical trial. |
| 18e | Data availability is reported. | Yes | Data Availability Statement | nuMoM2b data are available from the NICHD DASH repository upon application and approval. MMC data are not publicly available because of privacy regulations. |
| 18f | Analytical code availability is reported. | Partly | Data Availability Statement; Methods | Analytical procedures are described in the Methods. Public code release was not specified. |
| 19 | Patient and public involvement is reported if applicable. | Not applicable | — | No patient or public involvement was included in the design, conduct, reporting, or dissemination of this secondary analysis. |
| 20a | Participant flow is described, including number with and without outcome. | Yes | Section 3.1; Supplementary Figure S4 | Participant flow and sequential exclusions are reported in Section 3.1 and Supplementary Figure S4. |
| 20b | Participant characteristics and missing data are summarized. | Yes | Table 1; Supplementary Tables S1–S2, S6, and S9 | Baseline characteristics are shown in Table 1. Univariate associations are provided in Supplementary Tables S1 and S2. Included versus excluded pregnancies are compared in Supplementary Table S6. Predictor-level missingness is reported in Supplementary Table S9. |

| Item | Checklist item | Reported in manuscript | Location | Comments |
|---|---|---|---|---|
| 20c | Distribution of predictors and outcomes is compared between development and evaluation data if applicable. | Partly | Section 3.2; Supplementary Table S12 | Outcome prevalence is reported for both cohorts. The MMC cohort has been described previously [33]. |
| 21 | Number of participants and outcome events in each analysis is specified. | Yes | Sections 2.5, 2.6, and 3.1; Supplementary Tables S4, S5, S7, S8, S12, and S13 | The final analytic sample and event counts for primary and LGA sensitivity outcomes are reported. Event rates are also shown in model-performance and threshold-based operating-characteristic tables. |
| 22 | Full model specification or availability is provided to support future evaluation or implementation. | Partly | Section 2.5; Table 2; Figures 2–3 | The predictors in each model are listed in Table 2. Modeling procedures and feature contributions are reported. The study does not present a finalized deployable clinical model object. |
| 23a | Model performance estimates are reported with confidence intervals and relevant plots. | Yes | Section 3.2; Figure 1; Table 3; Supplementary Figures S1–S3 and S5; Supplementary Tables S4, S5, S7, S8, S10, S12, and S13 | Discrimination, precision–recall performance, calibration, threshold operating characteristics, non-linear classifier sensitivity comparisons, model-complexity analysis, the limited evaluation in the MMC cohort, and WHO versus INTERGROWTH-21st chart-comparator analyses are reported. |
| 23b | Heterogeneity in model performance across clusters is reported if examined. | Not performed | — | Center-or cluster-specific performance heterogeneity was not evaluated. |
| 24 | Model updating results are reported if performed. | Not applicable | — | No model updating after external validation was performed. |
| 25 | Overall interpretation of results is provided in context of objectives and previous studies. | Yes | Discussion, Sections 4.1–4.3 | The Discussion interprets the visit-updated prediction pattern, comparison with fetal growth chart–based benchmarks, dietary findings, clinical implications, and relationship to previous studies. |
| 26 | Limitations are discussed, including sample size, missing data, overfitting, representativeness, and generalizability. | Yes | Discussion, Section 4.4; Supplementary Tables S6 and S9 | Limitations include internal validation only, evaluation of the early-pregnancy model in the complete-follow-up cohort, selection bias, rare >4500 g outcome, |

| Item | Checklist item | Reported in manuscript | Location | Comments |
|---|---|---|---|---|
| | | | | missingness, ultrasound timing, Hadlock-derived EFW limitations, dietary recall bias, and need for external validation. Selection and missingness are further summarized in Supplementary Tables S6 and S9. |
| 27a | Handling of unavailable or poor-quality input data in future implementation is discussed. | Partly | Section 2.5; Discussion, Sections 4.3–4.4; Supplementary Table S9 | Missing data were handled during model development using k-nearest-neighbor imputation within cross-validation. Future implementation would require prospective assessment of input-data availability and quality. |
| 27b | Required user interaction and expertise for model use are discussed. | Partly | Discussion, Section 4.3 | The model is positioned as a potential adjunct for antenatal risk stratification rather than a replacement for clinical assessment. A specific implementation workflow was not evaluated. |
| 27c | Next steps for future research, applicability, and generalizability are discussed. | Yes | Discussion, Section 4.4; Conclusion | External validation and further evaluation before clinical implementation are explicitly stated. |

Note. TRIPOD+AI: Transparent Reporting of a multivariable prediction model for Individual Prognosis Or Diagnosis plus Artificial Intelligence. This checklist summarizes reporting coverage for the present model-development and internal-validation study. Items relating to external validation, model updating, deployment, or clinical implementation were marked as not applicable or partly reported where these analyses were not performed.

### Supplementary Table S12. Limited evaluation of a reduced early-pregnancy model in an independent Dutch cohort.

| Outcome | Model / evaluation setting | Predictors | Records (events, %) | AUROC (95% CI) |
|---|---|---|---|---|
| Birthweight > 4000 g | Model 1, internal validation in nuMoM2b | Full Model 1 predictor set (42 predictors) | 6,371 (505, 7.9%) | 0.628 (0.602–0.651) |
| Birthweight > 4000 g | Reduced early-pregnancy model, internal validation in nuMoM2b | Six harmonizable predictors | 6,371 (505, 7.9%) | 0.603 (0.577–0.627) |
| Birthweight > 4000 g | Reduced early-pregnancy model applied to MMC | Six harmonizable predictors | 12,043 (1,142, 9.5%) | 0.597 (0.580–0.613) |
| Birthweight > 4500 g | Model 1, internal validation in nuMoM2b | Full Model 1 predictor set (42 predictors) | 6,371 (77, 1.2%) | 0.721 (0.670–0.770) |
| Birthweight > 4500 g | Reduced early-pregnancy model, | Six harmonizable predictors | 6,371 (77, 1.2%) | 0.615 (0.556–0.678) |

| Outcome | Model / evaluation setting | Predictors | Records (events, %) | AUROC (95% CI) |
|---|---|---|---|---|
| | internal validation in nuMoM2b | | | |
| Birthweight > 4500 g | Reduced early-pregnancy model applied to MMC | Six harmonizable predictors | 12,043 (134, 1.1%) | 0.636 (0.589–0.681) |

Table note: The MMC cohort comprises retrospective electronic medical record data of pregnant women aged 18–45 years without preexisting diabetes who gave birth at Máxima Medical Center, Veldhoven, the Netherlands, between 2012 and 2017, and has been described previously [33]. The six harmonizable predictors were maternal age, pre-pregnancy BMI, and four race and ethnicity indicators (non-Hispanic White, non-Hispanic Black, Hispanic, and Asian/Other race or ethnicity). The reduced model was developed in nuMoM2b using the same modeling framework as the main models and was applied to the MMC data with the nuMoM2b coefficients and intercept unchanged, without refitting or recalibration. MMC records with missing predictor or outcome data were excluded. The MMC data included both nulliparous and multiparous women, and some women contributed more than one delivery record. This limited evaluation assessed discrimination only; calibration of absolute risks in MMC was not evaluated. 95% confidence intervals were obtained by bootstrap resampling at the level of women, with all delivery records of a woman resampled together. Periconceptional diet, most Visit-1 anthropometric measurements, and serial ultrasound measurements were not available in a comparable form in the MMC cohort. This analysis is a limited evaluation of a reduced predictor set and does not constitute external validation of the full visit-updated models. The Medical Ethics Review Committee of Máxima Medical Center reviewed the retrospective use of the MMC data and waived the requirement for formal ethical approval. AUROC: area under the receiver operating characteristic curve; BMI: body mass index; CI: confidence interval; MMC: Máxima Medical Center.

### Supplementary Table S13. Model complexity of the Combined model for birthweight >4500 g.

| Configuration | Number of predictors | Local approximate effective degrees of freedom | AUROC (95% CI) | ΔAUROC vs full model (95% CI) | Calibration slope | CITL | O/E |
|---|---|---|---|---|---|---|---|
| Full model | 65 | 24.2 | 0.821 (0.778–0.866) | — | 1.022 | −0.013 | 0.987 |
| 13-predictor model | 13 | 3.7 | 0.813 (0.764–0.859) | −0.008 (−0.037 to 0.018) | 1.056 | −0.011 | 0.990 |
| 10-predictor model | 10 | 3.0 | 0.803 (0.752–0.851) | −0.018 (−0.050 to 0.014) | 1.010 | −0.016 | 0.985 |

Table note: The analysis used 100 repetitions of stratified 10-fold cross-validation (6,371 pregnancies; 77 events). Within each outer training fold, predictors were ranked by their absolute standardized mean difference (Cohen's d), and the 13 or 10 highest-ranked predictors were retained; the validation fold was not used for ranking, and different folds could select different predictors. Within each outer training set, an inner 5-fold cross-validation was used to obtain out-of-fold scores, to which the Platt calibrator was fitted before application to the outer validation fold. The local approximate effective degrees of freedom and calibration measures reported here were obtained from a separate run of 100 repetitions. Calibration slope, CITL, and O/E were calculated from Platt-recalibrated out-of-fold probabilities averaged for each participant across those 100 repetitions. The local approximate effective degrees of freedom were calculated for the penalized model with the given set of active predictors and exclude the intercept; they do not account for the degrees of freedom used by predictor selection. Because predictors were selected within each outer training fold, the AUROCs of the reduced models evaluate the complete selection-and-fitting procedure. Out-of-fold predictions were averaged for each participant across the repetitions, and confidence intervals were obtained by bootstrap resampling of participants; differences in AUROC were estimated by paired bootstrap resampling. AUROC: area under the receiver operating characteristic curve; CITL: calibration-in-the-large; O/E: observed-to-expected ratio.
